# Associations Between Google Search Trends for Symptoms and COVID-19 Confirmed and Death Cases in the United States

**DOI:** 10.1101/2021.02.22.21252254

**Authors:** Mostafa Abbas, Thomas B. Morland, Eric S. Hall, Yasser EL-Manzalawy

## Abstract

We utilize functional data analysis techniques to investigate patterns of COVID-19 positivity and mortality in the US and their associations with Google search trends for COVID-19 related symptoms. Specifically, we represent state-level time series data for COVID-19 and Google search trends for symptoms as smoothed functional curves. Given these functional data, we explore the modes of variation in the data using functional principal component analysis (FPCA). We also apply functional clustering analysis to identify patterns of COVID-19 confirmed case and death trajectories across the US. Moreover, we quantify the associations between Google COVID-19 search trends for symptoms and COVID-19 confirmed case and death trajectories using dynamic correlation. Finally, we examine the dynamics of correlations for the top nine Google search trends of symptoms commonly associated with COVID-19 confirmed case and death trajectories. Our results reveal and characterize distinct patterns for COVID-19 spread and mortality across the US. The dynamics of these correlations suggest the feasibility of using Google queries to forecast COVID-19 cases and mortality for up to three weeks in advance. Our results and analysis framework set the stage for the development of predictive models for forecasting COVID-19 confirmed cases and deaths using historical data and Google search trends for nine symptoms associated with both outcomes.

## Background

In December 2019, an outbreak of the coronavirus disease (COVID-19) caused by the spread of the 2019 novel coronavirus (SARS-CoV-2) originated in the city of Wuhan in China. Due to the exponential worldwide spread of the virus, the World Health Organization (WHO) declared COVID-19 a pandemic on March 11, 2020. As of January 11, 2021, and according to John Hopkins University COVID-19 Dashboard (available at https://coronavirus.jhu.edu/map.html), the number of COVID-19 worldwide confirmed cases reached more than 90 million (including more than 22 million cases in the US) and the number of global deaths reached more than 1.9 million (∼376,000 in the US). To date, the COVID-19 pandemic has caused a huge negative global impact on the economy [1], health [2, 3], and education [4-6]), with the US as one of the top countries affected by this pandemic.

Extensive open access COVID-19 data, including test results, present outstanding opportunities for data scientists to apply a variety of statistical and machine learning approaches to characterize underlying factors governing variation in the pattern and rate of COVID-19 spread across the US. For example, Tang et al [7] applied functional principal component analysis (FPCA) to examine the modes of variation for COVID-19 confirmed case trajectories for 50 US states, quantified the correlations between confirmed case and death trajectories using functional canonical correlation analysis (FCCA), and grouped the 50 states into five subgroups where states shared similar COVID-19 spread patterns. Chen et al [8] used nonnegative matrix factorization (NMF) followed by a k-means clustering procedure applied to the NFM coefficients to cluster the COVID-19 confirmed case trajectories for 49 US states into 7 groups and investigated the dynamics of the clustering results over time. Similar analyses had been applied to various regions in the world. For instance, Carroll et al [9] used FPCA to investigated the modes of variation in COVID-19 case and death trajectories for 64 countries for an interval of 64 days and used functional regression analysis to show significant association between reduced workplace mobility and lower spread of the virus as well as association between baseline demographic (i.e., population density and percentage of population above 65 years) and doubling rates of mortality. Boschi et al [10] used a functional clustering approach based on probabilistic *k*-mean with local alignment [11] to investigate the patterns of COVID-19 mortality across 20 Italian regions and applied functional regression analysis [12] to show strong associations between COVID-19 mortality and both local mobility and positivity.

Recently, Google released the Google COVID-19 search trends symptoms dataset (version 1.0) [13]. The dataset provides daily (and weekly) time series of anonymized and aggregated searches for more than 400 medical symptoms at the state and county levels. The goal is to provide an open-access dataset to accelerate scientific and public health insights into the spread and impact of COVID-19 while maintaining the privacy of Google users. Traditional approaches for forecasting the spread of a virus (e.g., seasonal flu) infection were found to predict results with one or two weeks delay [14]. Since its presentation in November 2008, Google Flu Trends (GFT), an online tool that uses aggregated Google search queries data to provide daily estimates of the occurrence of flu two weeks in advance [15-17], had been shown to improve the predictive performance of traditional surveillance and forecasting systems [14, 18, 19]. Motivated by these results and several other studies demonstrating the viability of Google search trends in infectious disease surveillance and early epidemic prediction [20, 21], we hypothesize that Google search trends for COVID-19 related symptoms could be used in combination with historical COVID-19 data to predict future COVID-19 spread and mortality rates.

To the best of our knowledge, no models for forecasting COVID-19 spread or mortality using the Google COVID-19 search trends symptoms database have been developed yet. The major aim of this study is to establish the foundation for developing such prediction models via identifying relevant symptoms associated with COVID-19 confirmed cases and deaths. Additionally, we apply functional data analysis techniques to investigate modes of variation and patterns of COVID-19 positivity and mortality as well as Google search trends for selected COVID-19 related symptoms in the US.

## Methods

### Data

The numbers of daily COVID-19 confirmed cases and deaths were obtained from the Centers for Disease Control and Prevention (CDC) at https://data.cdc.gov/Case-Surveillance/United-States-COVID-19-Cases-and-Deaths-by-State-o/9mfq-cb36 (accessed on November 6th, 2020). The data were collected at the state-level including the 50 states as well as the District of Columbia (DC) (hereafter, the 51 US states).

The study period included the 245 days beginning March 1, 2020 (when the COVID-19 outbreak was declared a national emergency in the US) through October 31^st^, 2020. To account for variation in population size in each state, we normalized the number of cases to count per million people in each state using 2019 census population estimates.

In addition to the two COVID-19 time-series datasets collected from CDC, we experimented with 422 time-series datasets extracted from the Google COVID-19 Search Trends Symptoms dataset (in short, symptoms dataset) [13]. The symptoms dataset was released by Google on September 2^nd^, 2020 and is publicly available at: https://github.com/google-research/open-covid-19-data/. The dataset includes aggregated state-level daily Google searches for 422 symptoms and conditions that may relate to COVID-19. The data were normalized by scaling the daily symptom search count proportional to the total search activity in each state during the day. We also included data extracted from the 2019 US Census data (available at https://www.census.gov/data.html) for four risk factors [22-24] representing the percentage of the state-level population with: black race; age greater than 65 years; unemployment status; and income below the poverty level.

### Statistical analysis framework

In our analyses, we modeled the COVID-19 state-level data from CDC and Google search trends as functional data. Specifically, we transformed per-state time-series CDC and Google data collected during the study time interval into smoothed function curves (See Fig. 1 as an example). Using these curves as data atoms, functional data analysis (FDA) provides tools analogous to statistical and multivariate analysis tools where random variables are replaced with random functions [12]. We used functional principal component analysis (FPCA) [12, 25] to analyze variations in the data and detect patterns of the COVID-19 spread and mortality in the US. To quantify the similarity between two functional datasets (e.g., COVID-19 case trajectories and Google search trends for one of the COVID-19 related symptoms such as hypoxemia), we used two functional correlation analysis methods. First, we used the robust and relatively fast dynamic correlation procedure [26, 27] to quantify and rank the associations between COVID-19 trajectories and each of the 422 symptoms search trends. Second, we used the functional canonical correlation analysis (FCCA) method [12, 28] to depict the pairwise correlations between COVID-19 trajectories and each of the top nine symptoms search trends identified from the first step. In what follows, we provide brief summaries of the functional data analysis techniques used in this study.

**Fig. 1.**
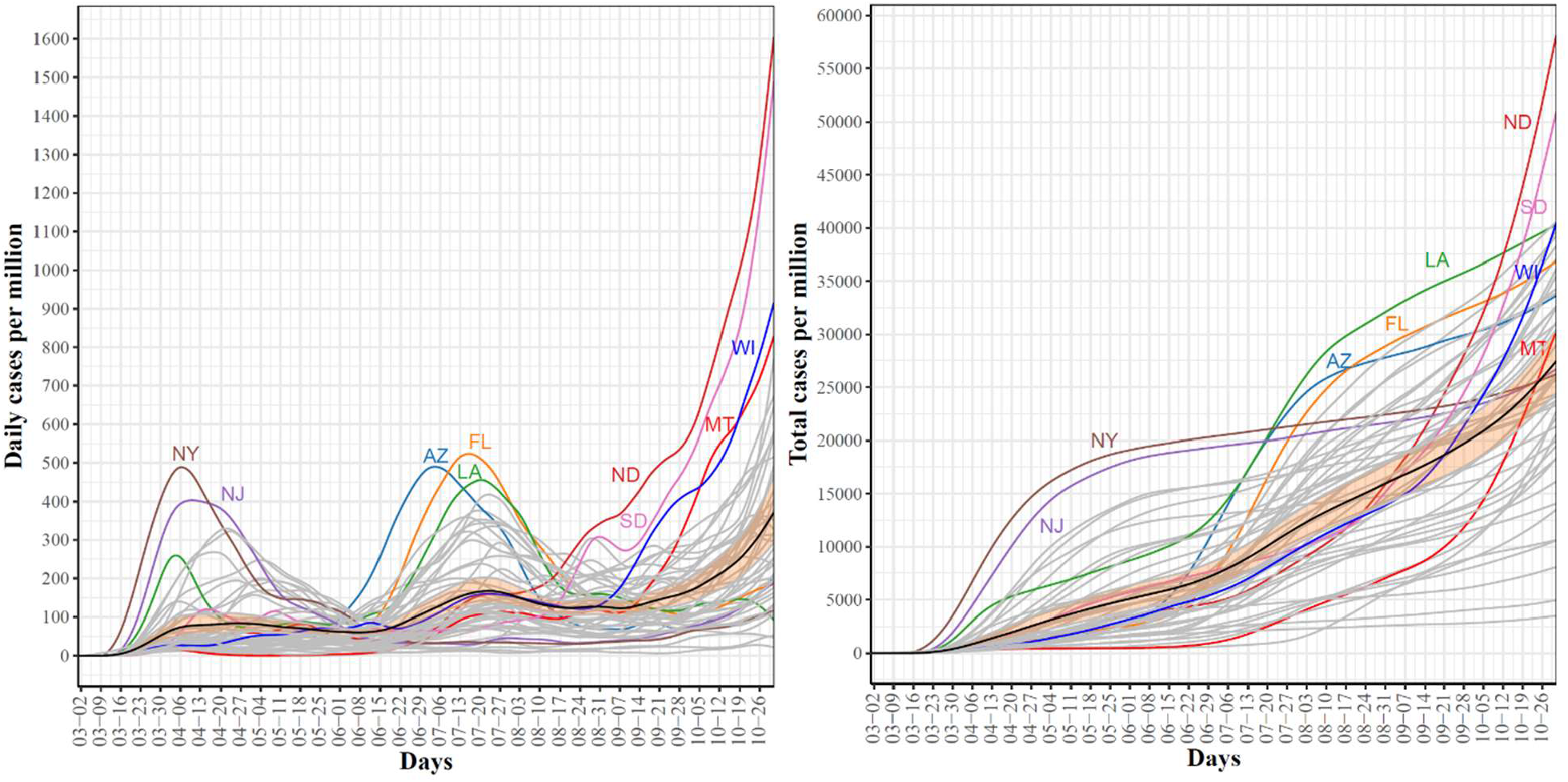
Trajectories for COVID-19 daily (left) and total (right) per million confirmed cases for the 51 US states. The mean curve is highlighted in black and the orange ribbon corresponds to the 95% confidence band.

### From time-series to functional data

Let {*t*_*ij*_: *i* = 1,2, …, *N and j* = 1,2, …, *T*} be an observed time-series dataset for N=51 states over T = 245 days. We transformed each time-series into a function using a weighted linear combination of *K* basis functions (i.e., 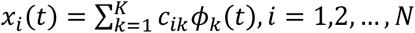 where *ϕ*_*k*_ is the *k*^*th*^ basis function). In our experiments, we used the cubic B-spline as the basis functions and set *K* equals to *T*. The coefficients *c*_*ik*_ where estimated using the roughness penalty [25, 29, 30] on the second derivative of each curve.

Using this data representation, we: i) examined the trajectories, and mean and variance curves to identify trends in the data; ii) applied several FDA approaches to the three functional datasets considered in this study.

### Functional principal component analysis

Functional principal component analysis (FPCA) is a powerful and widely used tool for linear dimensionality reduction of the functional data [12, 25]. In multivariate analysis (MVA), PCA is used to: i) reduce the number of (potentially) correlated variables to a smaller number of uncorrelated variables (principal components); ii) explore the latent relationships between variables. PCA determines the weights for combining the input variables into principal components with maximized variations. Specifically, the principal component scores *f*_*i*_ of a sample *x*_*i*_ are determined as 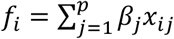, where the unknow weight vector *β*_*j*_= (*β*_*1*_, …, *β*_*P*_) is estimated using the following step-wise procedure [12]:

1. Determine the first eigenvector *ξ*_*1*_ that maximize 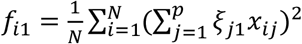 such that 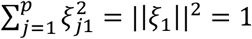
2. Repeat this step to compute subsequent eigenvectors *ξ*_m_for *m* = 2, …, *p*. On the *m*^*th*^ step, compute *ξ* _*m*_ such that 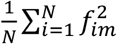 is maximized and subject to the following constraints

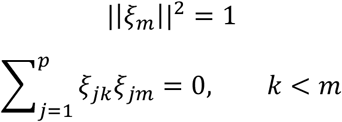

For functional data, the preceding procedure could be used with functions replacing vectors, continuous index *t* replacing the discrete index *j* and integrals replacing summations. For more details, interested readers are referred to [12].

For analyzing the variation in the functional data, we applied the concept of modes of variations for functional data [31] which visualizes the range of FPC scores. The *k*^th^ mode of variation is the set of functions computed as

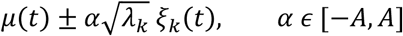

Where *μ*(*t*) is the mean curve and *λ*_*k*_ is the *k*^*th*^ eigenvalue and *A* = 2.

### Functional Clustering

An interesting application of FPCA is to represent each functional curve *x*_*i*_(*t*) using its FPCA scores in order to enable the application of traditional multivariate statistical and machine learning approaches. For clustering the functional curves of the 51 states, we represented each functional curve using the normalized first four FPCA. We then used the *k*-means algorithm [32, 33] and the Euclidean distance to cluster the data into *k* groups. We evaluated the clustering results for different choices of *k* (i.e., *k* = {2,3, …, 10}) using the majority rule applied to 30 different selection criteria (included in NbClust R package (version 3.0) [34]).

### Dynamic Correlation

To quantify the similarity between per-state curves for confirmed cases and deaths to counterparts’ curves for 422 symptoms, we used dynamic correlation [26, 27]. Dynamic correlation is a non-parametric functional data analysis technique developed to measure the correlation between two random functions at individual and population levels. Given two random functions, dynamic correlation can be viewed as the cosine similarity between them after population-centering [27]. It is worth noting that the name ‘dynamic correlation’ reflects the centering around a dynamic (time-varying) instead of a static population mean and the estimated correlation coefficient is a static value between -1 and 1. In our experiments, we used an implementation of the dynamic correlation methods proposed in [26] provided via the ‘dynCorr’ R package (version 1.1.0) [35].

### Functional Canonical Correlation Analysis

Canonical correlation analysis (CCA) [12, 28] quantifies the associations between two sets of variables by transforming both of them into a common lower dimensional space with maximum correlations. Given two random vectors *X* and *Y*, the CCA finds two weight vectors *u* and *v* such that the two linear transformations *u*^T^*X* and *v*^T^*Y*, also called the canonical variables, are maximally correlated. In 1993, Leurgans et al [36] adapted the CCA for functional data. Given two random functions, *X*(*t*) and *Y*(*t*), the functional correlation analysis (FCCA) seeks two weigh functions *u*(*t*) and *v*(*t*) such that 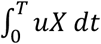 and 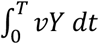, called functional correlation variables, are maximally correlated. In our analysis, we used the cca.fd function from the fda R package [37] to conduct FCCA with roughness penalties on the second derivative of each curve.

## Results

### Trajectories of COVID-19 confirmed and death cases in US

Fig. 1 shows the trajectories for COVID-19 daily (left) and cumulative (right) per million confirmed cases for the 51 US states. In the trajectories of the confirmed daily COVID-19 cases, the mean curve (black line) shows two local maxima (i.e., waves). The first wave peaked on April 6^th^ where the largest numbers of confirmed cases were reported in NY and NJ. The second wave peaked on July 20^th^ where the leading states were FL, AZ, and LA. The mean curve shows that another local maximum with larger numbers of cases is expected to be formed after October 31^st^ (end of our study time interval). In the 3^rd^ wave, ND and SD are expected to contribute the largest numbers of confirmed COVID-19 cases. The trajectories of total COVID-19 confirmed cases complement the trajectories of daily cases and reveal several interesting patterns. First, the two leading states in the first wave, NY and NJ, had total cumulative case rates greater than the mean from the start of the pandemic until the last week of October. However, their curves seem to have flattened since mid-June. Second, AZ and FL, the two leading states in the second wave, were below the mean curve until mid-June after which their curves were above the mean curve. The 3^rd^ leading state in the second wave, LA, followed a pattern that was different from AZ and FL. LA curve was always above the mean curve. Third, the leading states in the 3^rd^ wave had total numbers below the mean and started to cross the mean curve in the last week of August (the time when most schools and universities had opened for the new academic year). Fourth, by the end of our study time duration, the cumulative numbers of confirmed COVID-19 cases in the 3^rd^ wave leading states (ND, SD) exceeded the numbers for the 2^nd^ wave leading states (AZ, FL), which themselves exceeded the numbers for the leading states in the 1^st^ wave (NY, NJ). Overall, the trajectories of the confirmed COVID-19 cases demonstrated variations in the spread of COVID-19 across the US states.

Fig. 2 shows the trajectories for COVID-19 daily (left) and cumulative (right) per million death cases for the 51 states. For the daily trajectories, the mean curve had one global maximum which occurred in mid-April and starting mid-June the mean curve was almost horizontal until mid-October where the curve seemed to start going up again. The global maxima in the mean curve of daily death cases corresponds to the first wave of the COVID-19 spread in the US and therefore it is not surprising that most deaths are reported in NY and NJ, the two leading states in the first wave. Interestingly, the NJ curve of daily death cases had a unique pattern with two peaks occurring on April 20^th^ and June 25^th^. This peak reflects the sudden jump in the number of COVID-19 deaths in NJ when 1854 probable COVID-19 deaths were added. The trajectories of the total COVID-19 death cases show that from the starting date of our study period until June 25^th^, NY had the largest cumulative number of COVID-19 deaths. After June 25^th^ and until the end of the study period, NJ is the leading state in terms of the total per million number of COVID-19 deaths. At the end date of our study period, the top leading states in terms of total per million number of deaths are NJ, NY, MA, CT, LA, and RI.

**Fig. 2.**
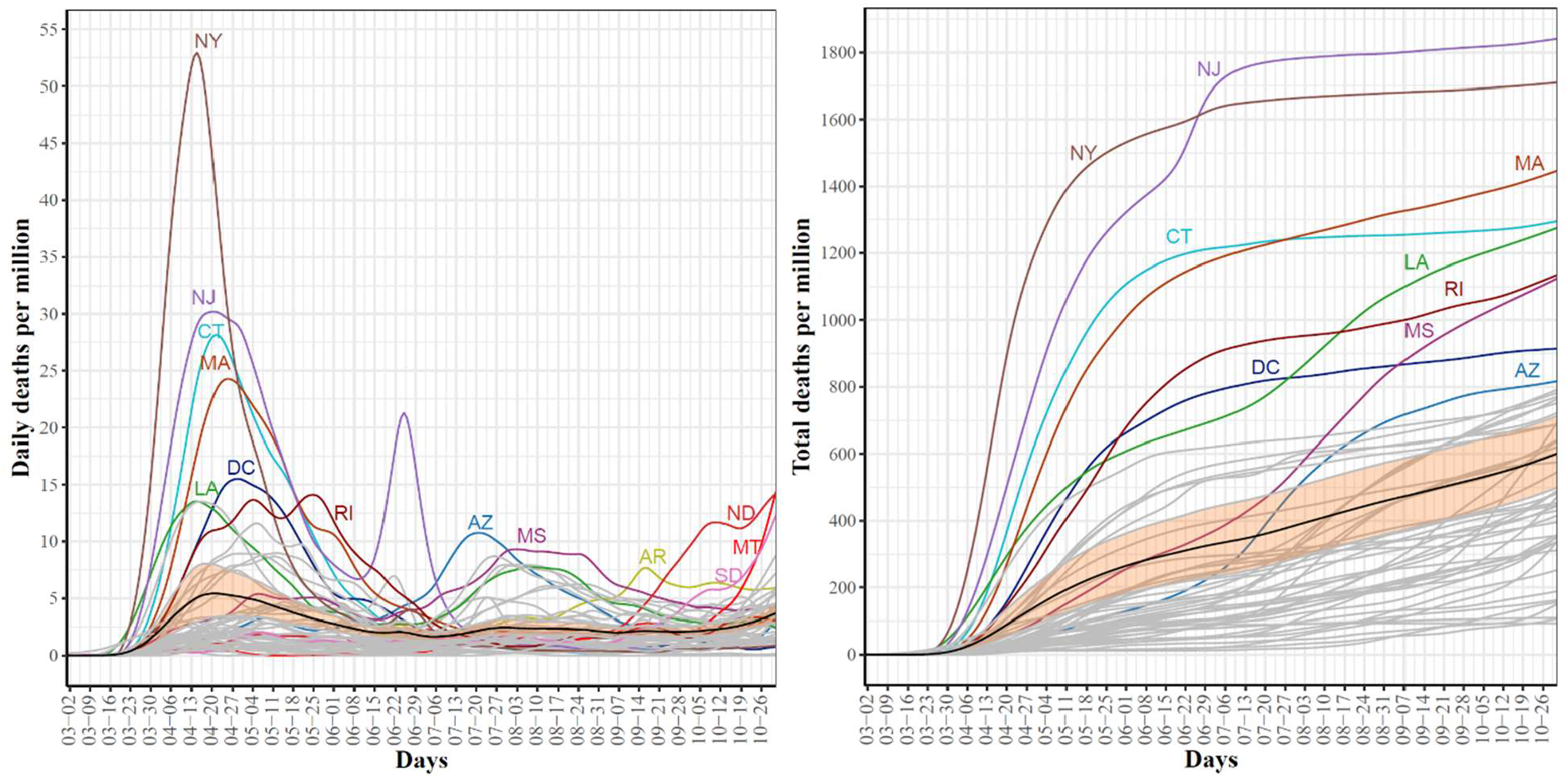
Trajectories for COVID-19 daily (left) and total (right) per million death cases for the 51 US states. The mean curve is highlighted in black and the orange ribbon corresponds to the 95% confidence band.

### Modes of variations in COVID-19 confirmed and death cases trajectories

To further explore the variations in the curves representing COVID-19 daily confirmed and death cases, we utilized the FPCA to project the data in a lower dimensional space. Fig. 3 shows the top four eigenfunctions counting for 95.28% of the variations in the curves shown in Fig. 1. The first eigenfunction *ξ*_1_ was less than zero until June 4^th^ and then seemed to be exponentially increasing. This suggests the variability between states started to increase in early June and as we move forward the variability increases more. The second eigenfunction *ξ*_2_ had two peaks. The first peak was negative and was observed in mid-April (close to the time of the first wave peak). The second peak was positive and occurred in mid-July (close to the time of the peak in the second wave). The curve also shows that a third negative peak is about to be formed likely in a future time that is close to the time of the peak in the third wave. The third eigenfunction *ξ*_3_ was always positive (except during the first 10 days of March) and had a global maximum in mid-April and another local maximum in mid-July. The fourth eigenfunction *ξ*_4_ was sinusoidal with alternating negative and positive peaks occurring in mid-April, mid-May, end of June, and end of August. Since the top two eigenfunctions accounted for 82.7% of the total variation in the data, we projected the 51 states in a two-dimensional space determined by the first two FPC scores. This projection of the 51 curves is shown in Fig. 4. We found that the first wave leading states, NY and NJ, had negative FPC1 and lowest FPC2 scores. The three leading states in the second wave (FL, AZ, and LA) had negative FPC1 scores and the highest FPC2 scores. Finally, the leading states in the third wave (e.g., ND, SD, and WI) had the highest FPC1 scores but negative FPC2 scores. Fig. 5 shows the spectrum of curves representing the range of FPC1 and FPC2 scores. The first mode of variation captured greater variability in the data in September and October and showed that from mid-May to end of June, all curves were close to the mean curve. The top leading state in each of the three waves (NY, AZ, and ND) could be easily identified as outliers from the spectrum of curves for FPC1 scores. The second mode of variation captured the shape of the curves and shows that the greatest variability among curves occurred during the second wave. Two inflection points were noted at the first of June and October, respectively.

**Fig. 3.**
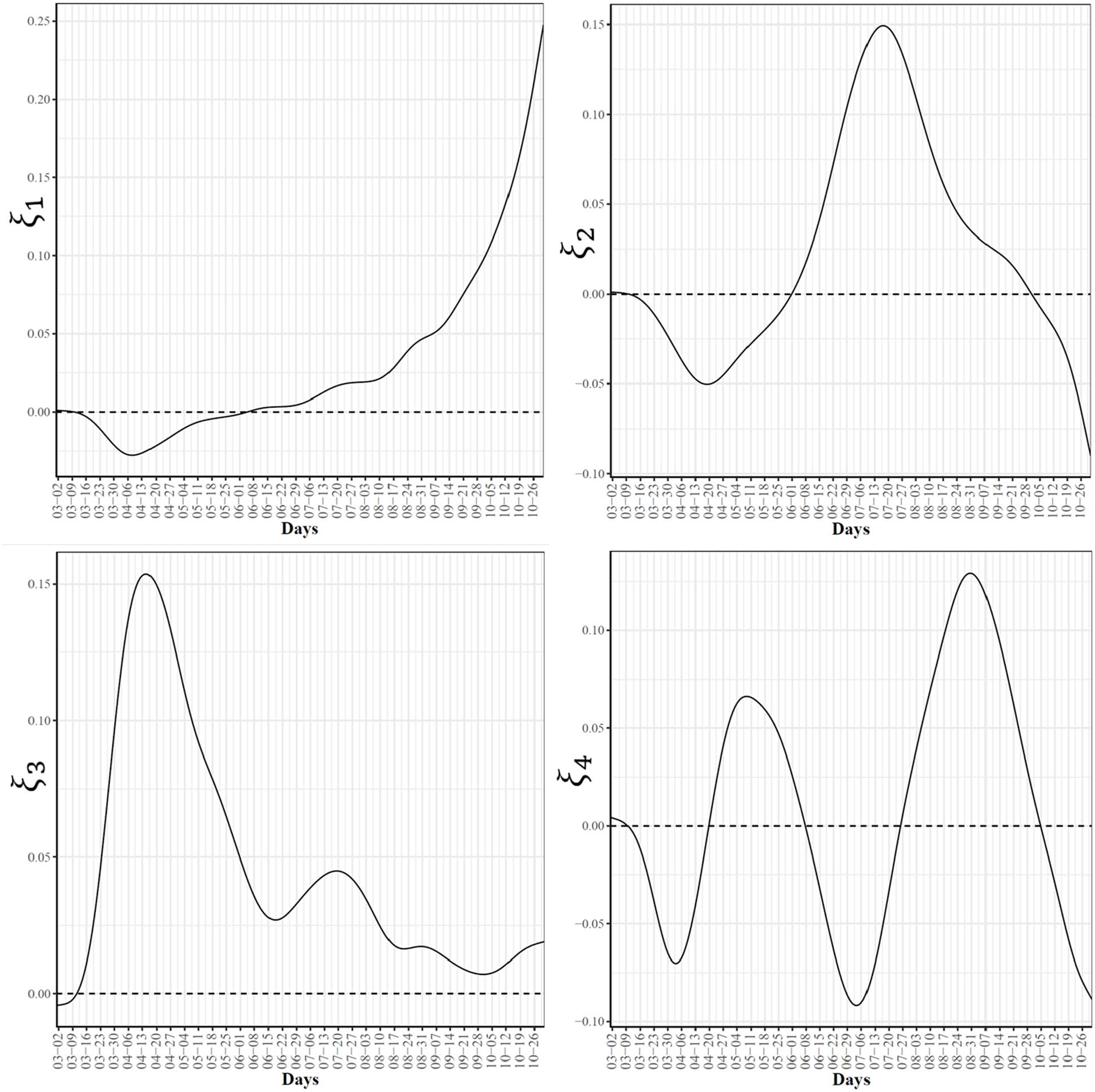
First four eigenfunctions of COVID-19 confirmed case trajectories.

**Fig. 4.**
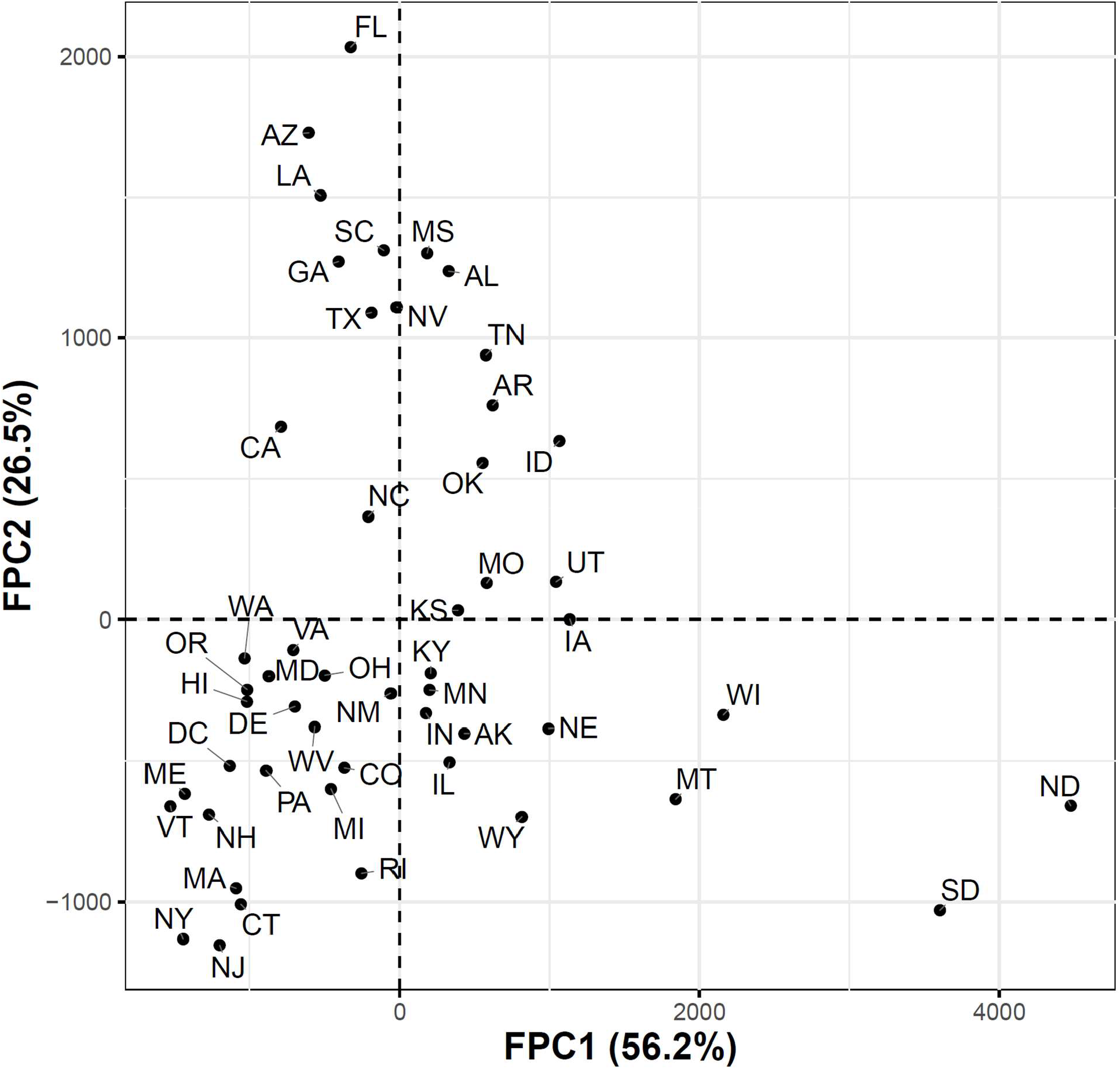
Scatter plot of the 51 US states in a two-dimensional space defined by the first two functional principal component scores of the COVID-19 case trajectories.

**Fig. 5.**
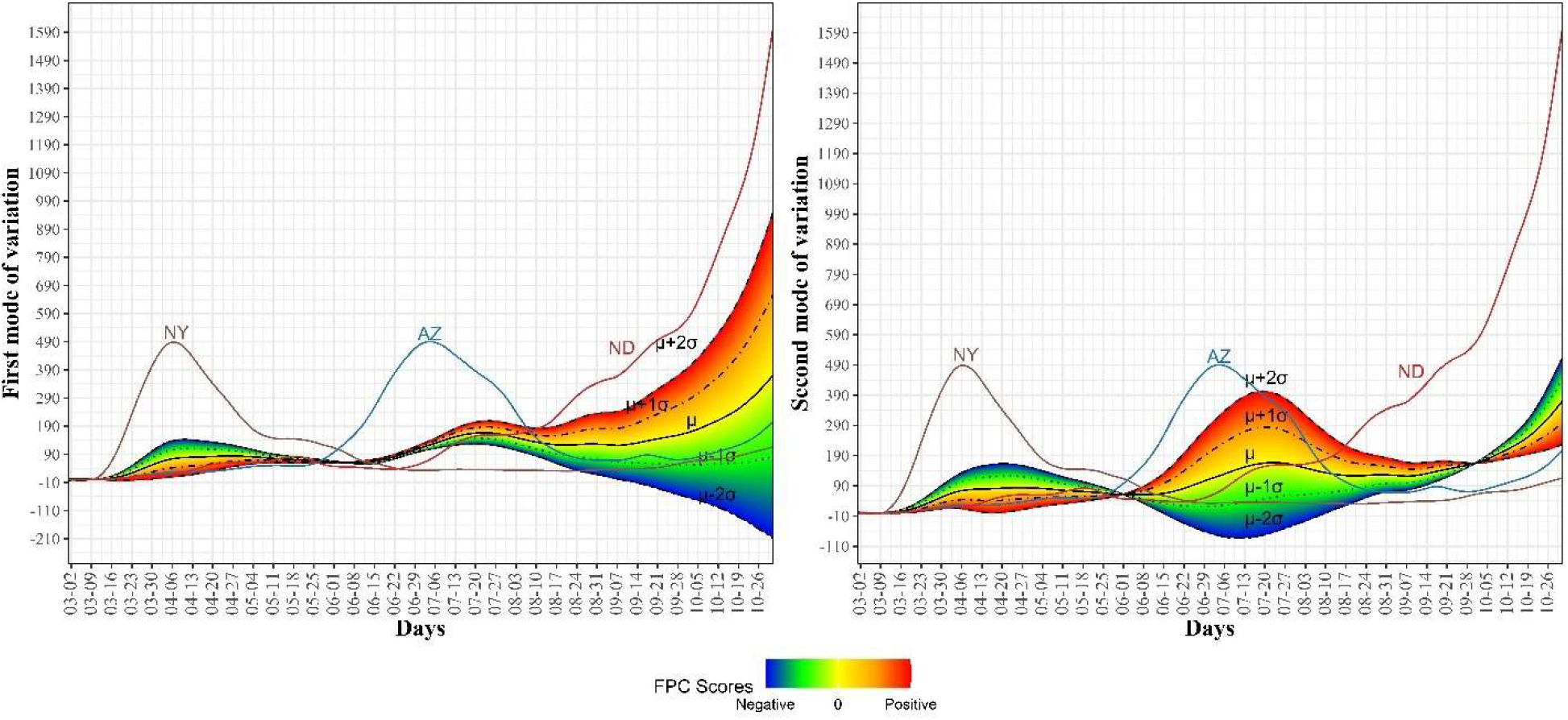
First and second modes of variations for the COVID-19 confirmed trajectories. The sample standard deviations of the first and second FPC scores are 1179.94 and 810.53, respectively. The curve for the leading state in each wave is also shown.

To examine the modes of variation in the COVID-19 daily death cases, we report the top four eigenfunctions (Fig. 6) as well as the two-dimensional representation of the 51 US states in the space defined by the first two FPC scores (Fig. 7). In Fig. 6, we noted that the first eigenfunction *ξ*_1_ had two positive peaks corresponding to the two peaks in the trajectories of NY and NJ, respectively (see Fig. 2). We also noted that the second eigenfunction *ξ*_2_ had a negative peak in mid_April and two positive peaks in mid-May and late July. These two eigenfunctions explained 83.8% of the variability in the COVID-19 daily death trajectories. Fig. 7 shows the 51 in the two-dimensional FPC space. Most of the states seemed to form a single cluster and outlier states represented the states with high rates of COVID-19 mortality. Although NY and NJ were two clear outliers, they were not close to each other in the FPC space which reflects the different patterns in the daily death trajectories of the two states (See Fig. 2). Fig. 8 shows the first and second modes of variations for COVID-19 death trajectories. The first mode of variation shows that the greatest variation in the data occurred in the third week of April. Starting mid-July, the variability in the data was consistently low. The second mode of variation seems to modulate the shapes of the curves and the inflection point at the third week of April corresponds to the time when the death rates in the first wave started to drop. In both modes of variation, the top leading state in each of the three waves (NY, AZ, and ND) appeared as clear outliers.

**Fig. 6.**
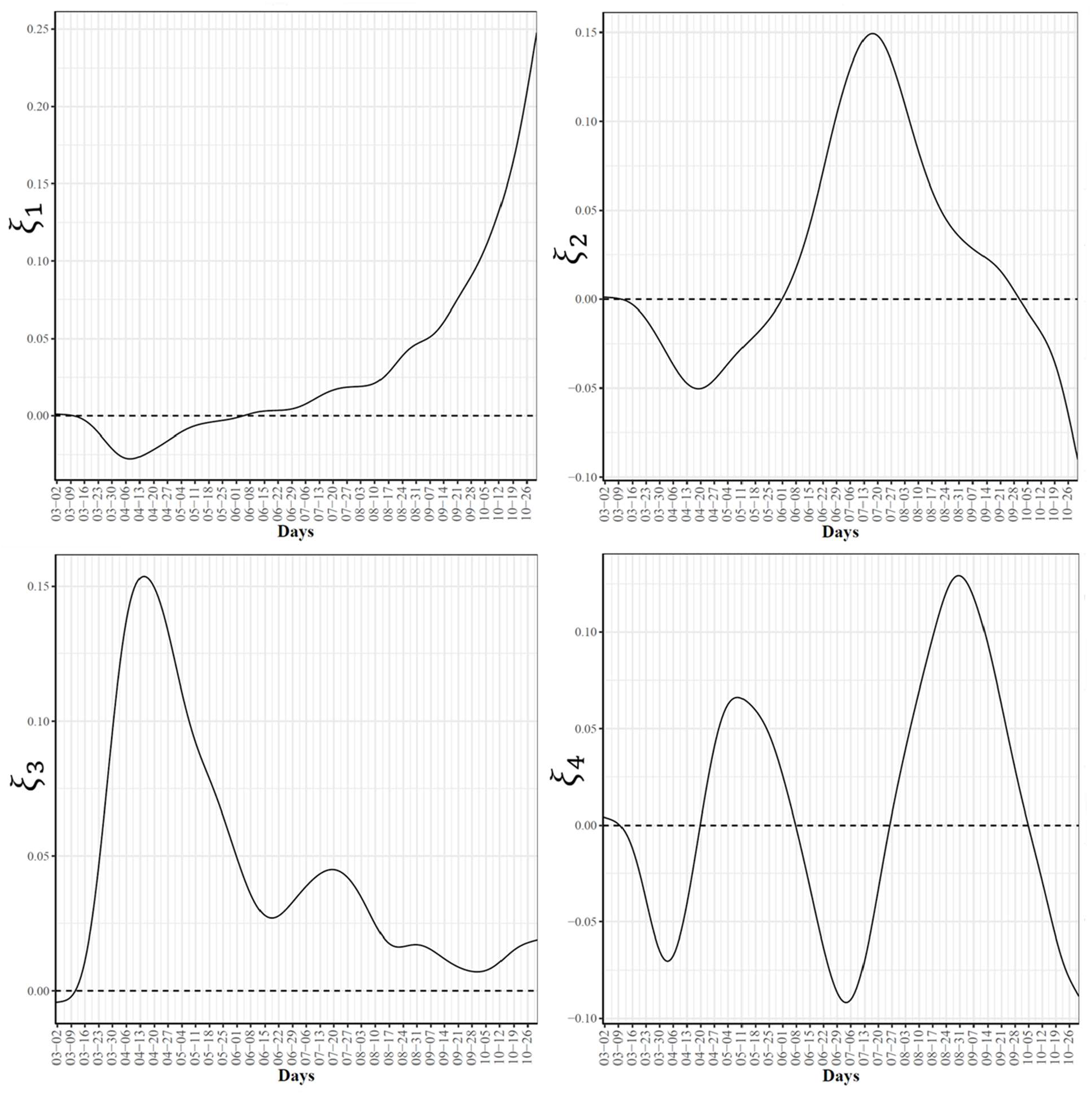
First four eigenfunctions of COVID-19 death trajectories.

**Fig. 7.**
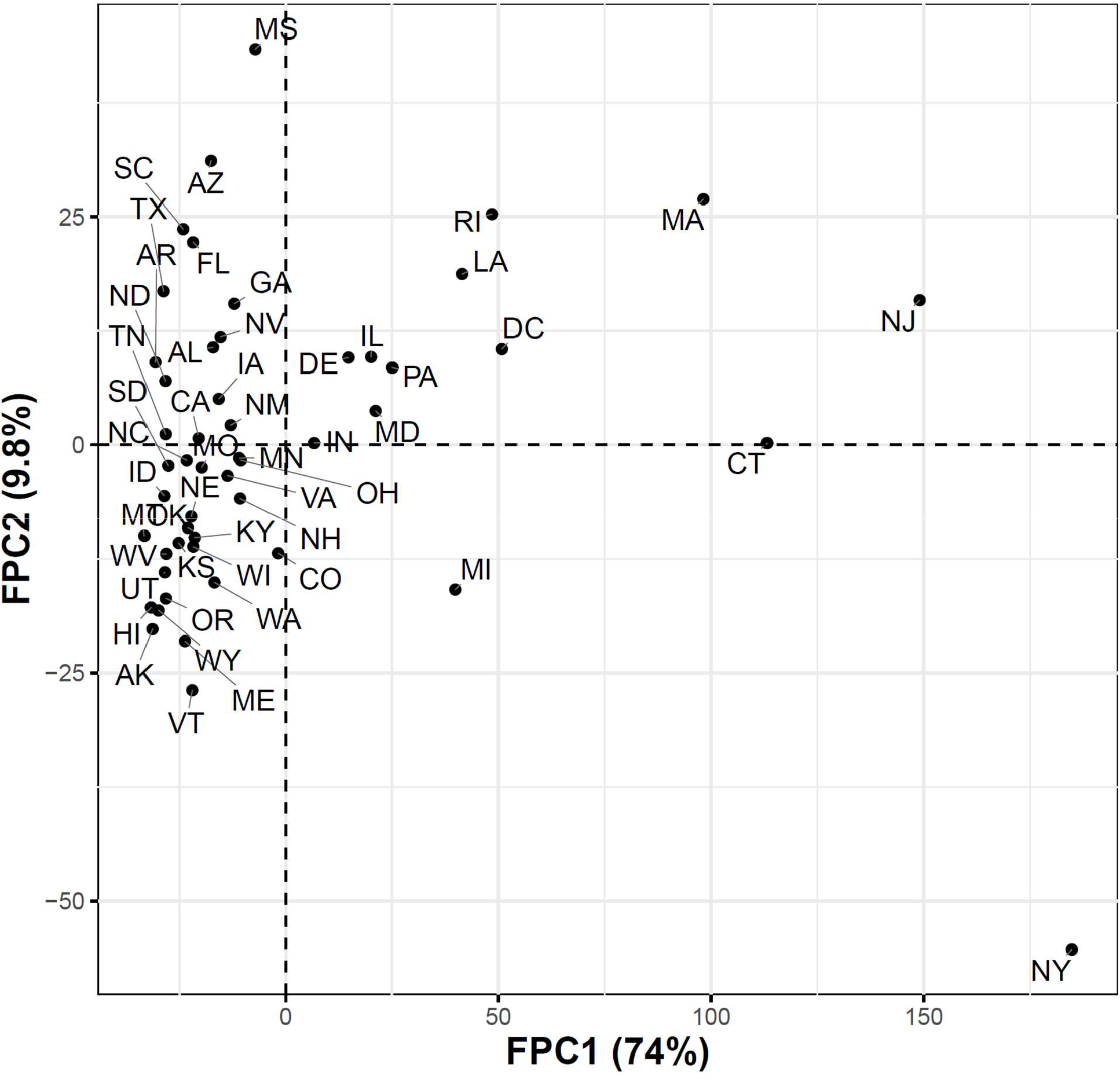
Scatter plot of the 51 US states in a two-dimensional space defined by the first two functional principal component scores for the COVID-19 death trajectories.

**Fig. 8.**
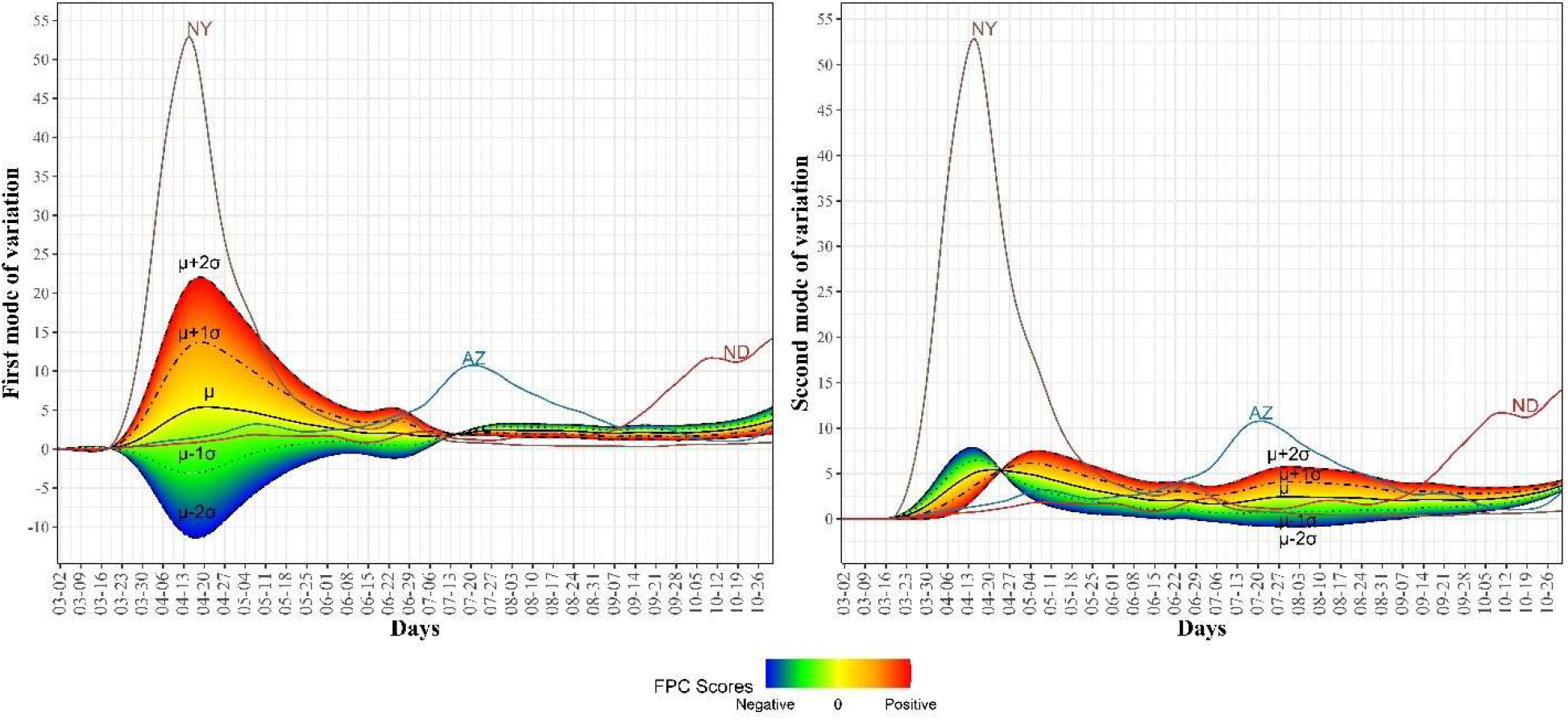
First and second modes of variations for the COVID-19 death trajectories. The sample standard deviations of the first and second FPC scores are 46.57 and 16.99, respectively. The curve for the leading state in each wave is also shown.

### Clustering US states by trajectories of COVID-19 confirmed cases

Fig. 9 shows the clustering of the 51 US states based on the trajectories of COVID-19 daily confirmed cases. We found that the optimal number of clusters was seven. This large number of clusters reflects the great variations in the spread of COVID-19 across different US states due to differences in states related to demographics of its population and health policies adopted by the US states.

**Fig. 9.**
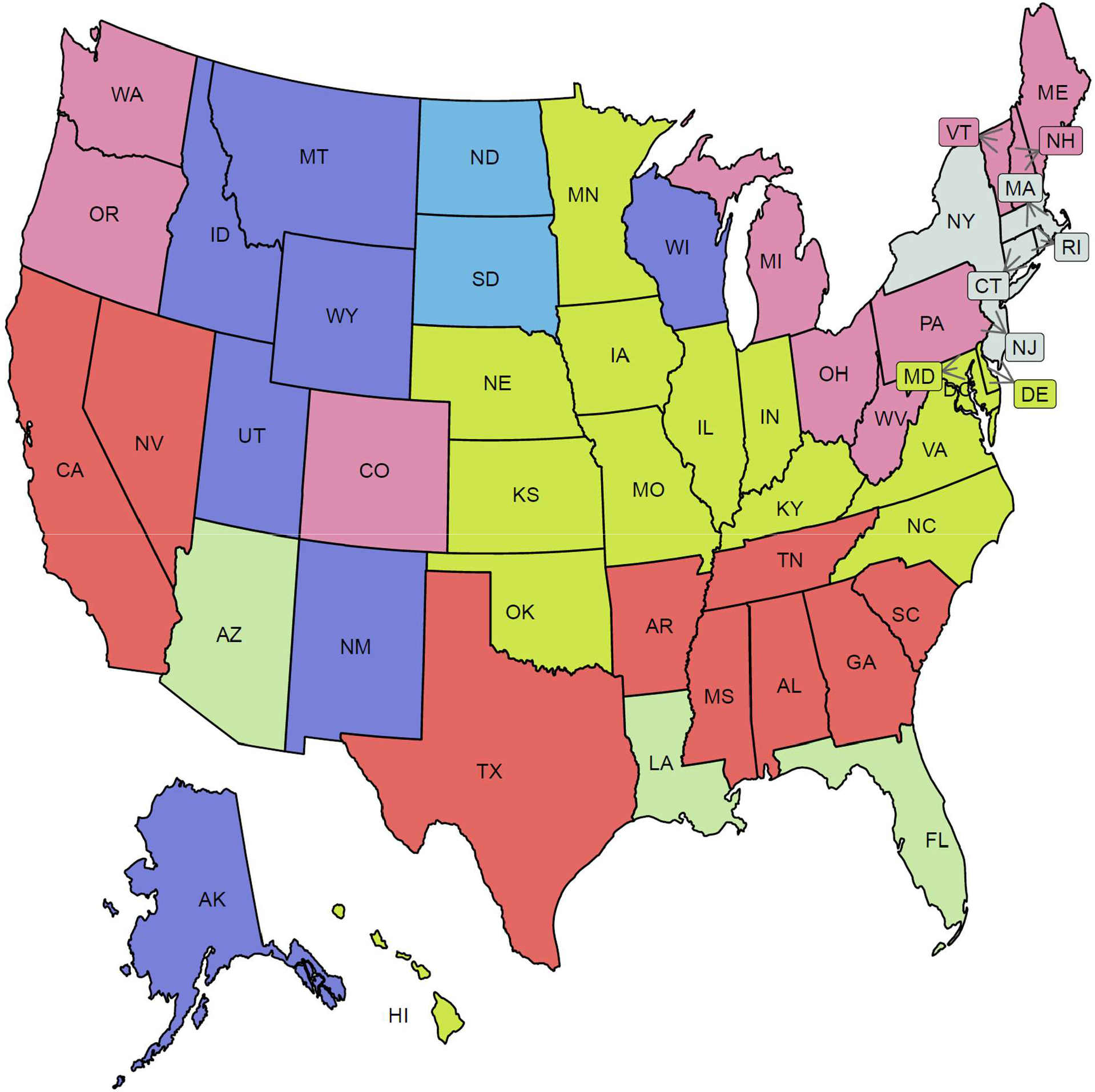
Clustering result of the 51 states based on the COVID-19 confirmed case trajectories. States assigned to the same cluster share the same color.

Fig. 10 characterizes the member states in each of the seven identified groups from the curves of COVID-19 confirmed cases. Cluster C1 included the three leading states in the second wave AZ, FL, and LA. Interestingly, none of these three states had shared borders with the other two states. States in this group encountered two peaks in the numbers of COVID-19 cases. The first peak (with a mean of 100 cases per million) occurred in the first week of June while the second peak (with a mean of ∼460 cases per million) occurred in mid-July. At the end of the study interval, another peak is about to be constructed with a mean of at least 150 cases per million. Cluster C2 included 10 states (CO, ME, MI, NH, OH, OR, PA, VT, WA, WV). This class covered states from North East, Mid-Atlantic, North West, and one isolated state, CO, from the Mid-West. Like the cluster C1 mean curve, the cluster C2 mean curve had two peaks which occurred in early June and 20^th^ of July but the number of cases in these two peaks was around 50-60 cases per million. Cluster C3 included six western states (AK, ID, MT, NM, UT, WY) and one Mid-West state, WI. The mean curve of cluster C3 had only one peak (with 150 cases per million) that occurred on the 20^th^ of July and a new peak (with more than 600 cases per million) will be formed after the end of the study duration. Cluster C4 had seven southern states (AL, AR, GA, MS, SC, TN, TX) and two western states (CA and NV). Cluster C4 had a mean curve with a dramatic increase in the number of cases started in the third week of May and reached a peak of 320 cases per million on the 20^th^ of July. The future expected peak is expected to have more than 250 cases. Cluster C5 had seven states (CT, DC, MA, NJ, NY, RI) which covers the two leading states in the first wave and five neighboring states from the North East and Mid-Atlantic regions. States of this cluster had an early peak in the third week of April with the average number of cases reaching 300. Then, the average number of cases dropped to less than 50 in mid-June before crossing 100 cases per day in mid-October. Cluster C6 had 14 states from the Mid-West and Mid-Atlantic regions (DE, HI, IA, IL, IN, KS, KY, MD, MN, MO, NC, NE, OK, VA). The mean curve of states in cluster C6 had three local maxima with 100, 150, 150 cases, respectively. The average number of cases exceeded 350 daily cases at the end of our study period. Cluster C7 included two states, ND and SD, which are the top two leading states in the third wave. The mean curve of these two states crossed the 100 daily cases for the first time in mid-July and crossed the 500 daily cases in the last week of September and reached more than 1500 daily cases by the end of October.

**Fig. 10.**
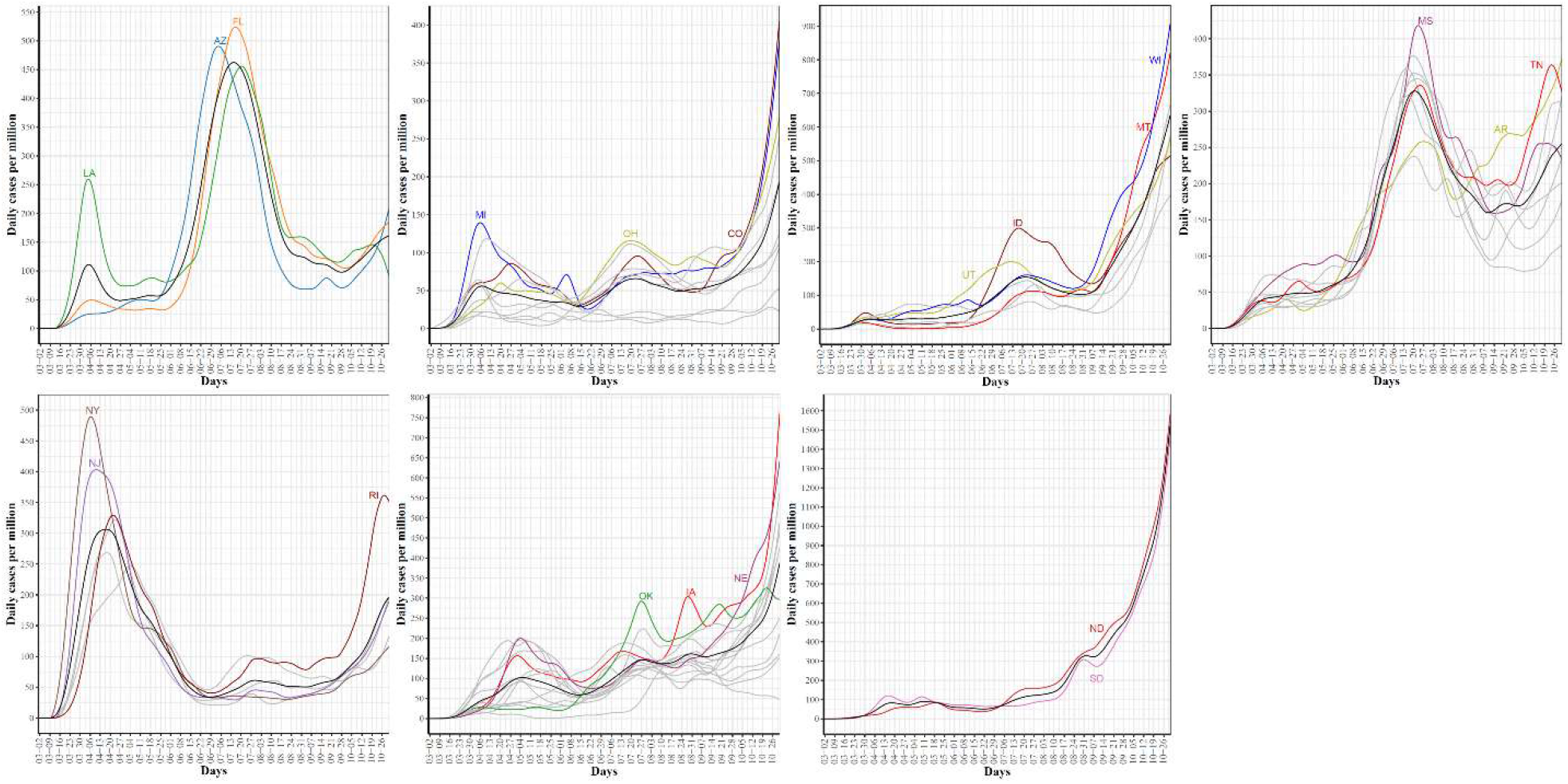
COVID-19 confirmed case trajectories of the states in each cluster. The mean curve for the states in each cluster is highlighted in black.

We also used four potential risk factors from 2019 census data to characterize the seven groups. Fig. S1 shows the boxplots of the seven groups for each risk factor. Table 1 summarizes these results using the median and the rank of these risk factors in each group.

**Table 1:** **Characterization (in terms of median and rank) of the seven groups of COVID-19 spread patterns across the US 51 states using four risk factors from 2019 US census data**.

| Census Variable | C1 | C2 | C3 | C4 | C5 | C6 | C7 |
| --- | --- | --- | --- | --- | --- | --- | --- |
| Black race | 16.9(2) | 4(5) | 1.5(7) | 17.1(1) | 13.6(3) | 9.2(4) | 2.9(6) |
| Age 65+ | 18.2(2) | 18.5(1) | 17.1(3) | 16.5(5) | 17(4) | 16.2(7) | 16.3(6) |
| Unemployment | 5(1) | 4.5(3) | 3.5(6) | 4.8(2) | 4.5(3) | 4(5) | 2.7(7) |
| Below poverty level | 13.5(2) | 11.2(5) | 10.4(6) | 13.8(1) | 10.4(6) | 11.4(3) | 11.3(4) |

### Clustering US states by trajectories of COVID-19 death cases

Clustering the 51 US states based on the trajectories of COVID-19 daily death cases yielded five clusters (See Fig. 11). Fig. 12 characterizes the member states in each group. Cluster C1 included nine southern states (AL, AZ, FL, GA, LA, MS, NV, SC, TX). Its mean curve showed two peaks that were observed in mid-April and last week of July with the average number of per million deaths equal 3 and 7, respectively. Cluster C2 included nine states (CT, DC, DE, IL, MA, MD, NJ, PA, RI). IL is the only member state in C2 that does not have any shared borders with any other member states in C2. The mean curve of C2 reached its global maximum in the last week of April with 15 per million deaths. Cluster C3 had 27 states (AK, CA, CO, HI, IA, ID, IN, KS, KY, ME, MI, MN, NC, NE, NH, NM, OH, OK, OR, TN, UT, VA, VT, WA, WI, WV, WY) and the average number of per million deaths for this cluster was always less than or equal 3. Cluster C4 included five states spanning North-West (MT), Mid-West (MO, ND, SD), and South-East (AR) regions. Its mean curve demonstrated that this cluster had the largest number of per million deaths since the beginning of October 2020. Cluster C5 had only one state, NY. The trajectory curve for NY followed the same pattern as the trajectory curves in C2 however NY reached a peak of 52 per million death cases in mid-April while the leading state in C2, NJ, reached a peak of 30 per million death cases. Table 2 characterizes each cluster using the median of four state-level risk factors from 2019 US census data. Interestingly, cluster C1 is ranked at the top for three risk factors (race is black, unemployment, and poverty), while it had the lowest rank for the percentage of the population older than 65 years.

**Table 2:** **Characterization (in terms of median and rank) of the seven groups of COVID-19 death patterns across the US 51 states using four risk factors from 2019 US census data**.

| Census Variable | C1 | C2 | C3 | C4 | C5 |
| --- | --- | --- | --- | --- | --- |
| Black race | 26.8(1) | 14.6(3) | 4.6(4) | 3.4(5) | 17.6(2) |
| Age 65+ | 16.5(5) | 17.1(2) | 16.8(4) | 17.2(1) | 16.9(3) |
| Unemployment | 4.9(1) | 4.6(2) | 3.9(4) | 3.8(5) | 4.4(3) |
| Below poverty level | 13.6(1) | 10.8(5) | 11.2(4) | 12.6(3) | 13(2) |

**Fig. 11.**
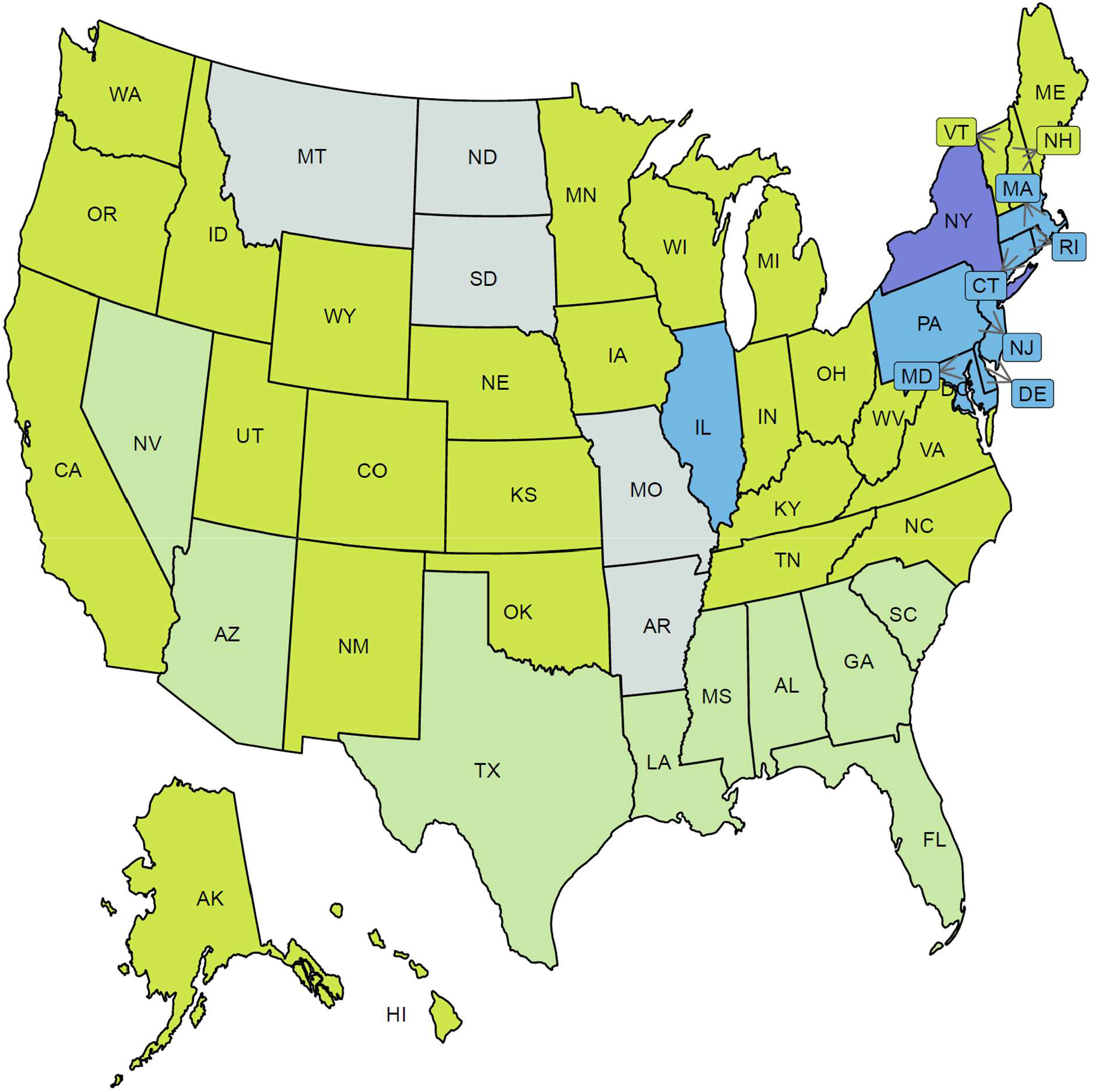
Clustering result of the 51 states based on the COVID-19 death trajectories. States assigned to the same cluster share the same color.

**Fig. 12.**
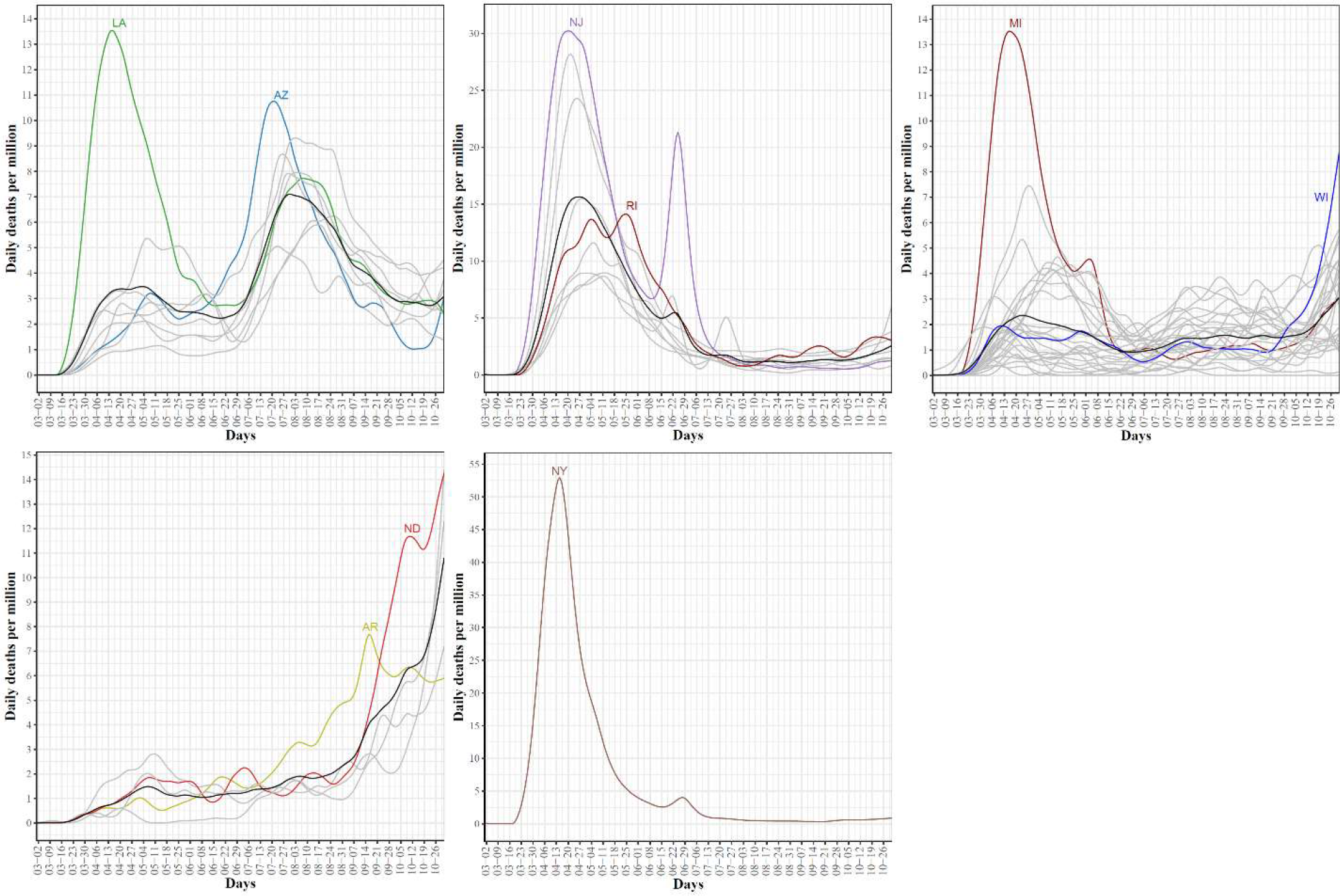
COVID-19 death trajectories of the states in each cluster. The mean curve for the states in each cluster is highlighted in black.

### Associations between Google COVID-19 search trends for symptoms and COVID-19 confirmed and death cases

Anonymized and aggregated state-level Google COVID-19 daily searches for symptoms were smoothed and treated as 422 functional datasets, one dataset for each symptom. Pairwise associations between each symptom functional dataset and functional datasets for COVID-19 confirmed and death cases, respectively, were quantified using dynamic correlation [26]. The complete results are reported in Tables S1 and S2. Table 3 summarizes the top 15 symptoms, sorted by the absolute value of the dynamic correlation estimates, associated with COVID-19 confirmed and death cases, respectively. We noted that the top 15 symptoms associated with COVID-19 daily confirmed cases had relatively higher dynamic correlations compared to the dynamic correlation estimates for the top 15 symptoms associated with COVID-19 daily death cases. Interestingly, hypoxemia, ageusia, and anosmia are amongst the top 5 symptoms in both cases. The table includes four symptoms with no clear relation to COVID-19: xeroderma, bruise, nodule, and genital wart. Interestingly, the four symptoms are related to dermatologic conditions.

**Table 3:** **Top 15 symptoms associated with COVID-19 confirmed cases and deaths, respectively. The nine symptoms shared in the two sets are highlighted in bold**.

| Symptom | Correlation | Symptom | Correlation |
| --- | --- | --- | --- |
| **Hypoxemia** | 0.78 | **Hypoxemia** | 0.65 |
| **Ageusia** | 0.69 | **Hypoxia** | 0.54 |
| **Anosmia** | 0.69 | **Pneumonia** | 0.48 |
| **Dysgeusia** | 0.65 | **Anosmia** | 0.46 |
| **Hypoxia** | 0.64 | **Ageusia** | 0.41 |
| Sinusitis | 0.62 | Panic attack | 0.40 |
| **Fever** | 0.61 | **Fever** | 0.40 |
| Low grade fever | 0.61 | **Shortness of breath** | 0.40 |
| Xeroderma | 0.56 | Chest pain | 0.40 |
| **Pneumonia** | 0.55 | **Dysgeusia** | 0.39 |
| **Chills** | 0.54 | Nodule | -0.38 |
| **Shortness of breath** | 0.54 | Genital wart | -0.37 |
| Cough | 0.54 | Bradycardia | 0.36 |
| Nasal congestion | 0.52 | Bronchitis | 0.35 |
| Bruise | -0.52 | **Chills** | 0.35 |

Fig. 13 shows the mean curves for COVID-19 daily confirmed and death cases as well as the mean curve with the top 3 symptoms associated with COVID-19 daily confirmed and death functional data, respectively. It should be noted that the correlations reported in Table 3 were not computed using the mean curves but using the state-level curves in each group. However, the mean curves summarized the trend in each group and showed interesting trends in the search curves that are correlated with the times of the three waves. For example, the mean curve for hypoxemia showed a drop in searches for this symptom in two time-intervals, 05/20 – 06/20 and 07/29 – 09/14, where a drop in the average number of daily confirmed cases is noticed in these two time-intervals. We also noted that the mean curves for ageusia and anosmia seemed to have the same trend. The dynamic correlation coefficients between COVID-19 daily confirmed curves and the curves for these symptoms was 0.69 in both cases.

**Fig. 13.**
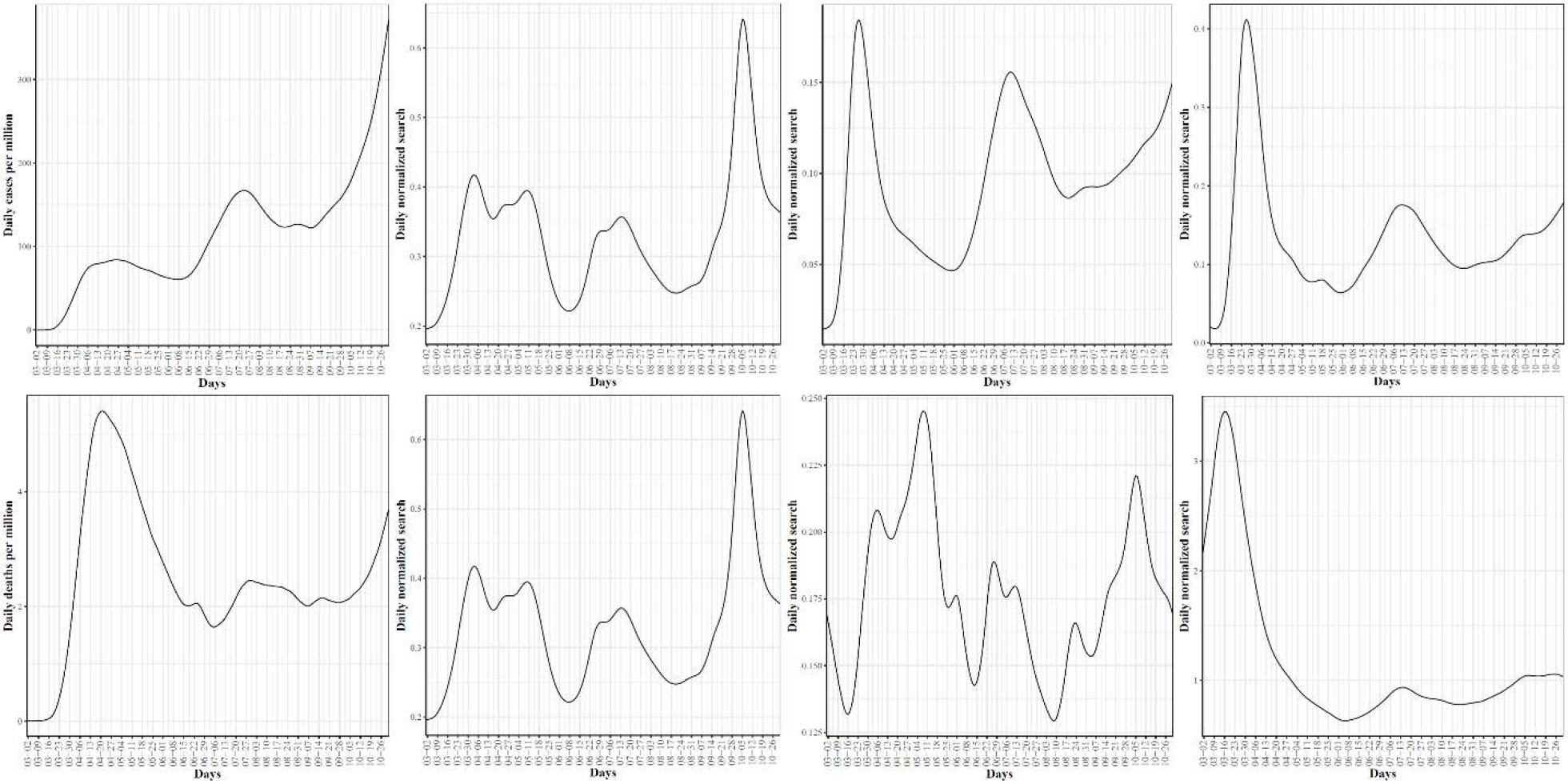
At the top are the mean curves for the COVID-19 confirmed cases, hypoxemia, ageusia, and anosmia searches, respectively. At the bottom are the mean curves for the COVID-19 death cases, hypoxemia, hypoxia, and pneumonia searches, respectively.

### Dynamics of associations between top Google search trends and future COVID-19 confirmed and death cases

Let 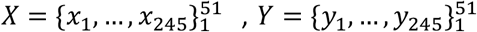 represents the time-series data for COVID-19 confirmed cases and Google searches for hypoxemia, respectively. As shown in Table 3, the dynamic correlation coefficient between these two functional datasets is 0.78. This high correlation score indicates a strong association between the number of searches for hypoxemia *y*_*t*_ and the number of COVID-19 cases *x*_*t*_ at any timepoint *t*. Additionally, we are interested in the correlation between *y*_*t*_and *x*_*t*+*k*_, *k* ≥ 1, which represents the association between the number of searches for hypoxemia in day *t* and the number of COVID-19 cases observed in the future (i.e., *k* days after *t*). Fig. 14 reports such dynamic correlations for the nine symptoms that are shared in the top associated 15 symptoms with COVID-19 confirmed (left) and death (right) cases for *k* = 0,1, …, 30 days. For correlations with COVID-19 confirmed cases, the highest correlations were observed at *k* = 0 for hypoxemia, hypoxia, and pneumonia, while for the remaining six symptoms the highest correlations (which are slightly greater than the correlations at *k* = 0) were noted for *k* ∈ [9 − 15]. The only feature that had a non-slight increase in its correlation coefficient compared to its initial correlation was “shortness of breath”. Correlation curves for hypoxemia, hypoxia, and pneumonia started to drop earlier than the curves for the remaining symptoms. At *k* = 30 hypoxemia and pneumonia had the lowest dynamic correlation coefficients compared with the correlations for the remaining symptoms.

**Fig. 14.**
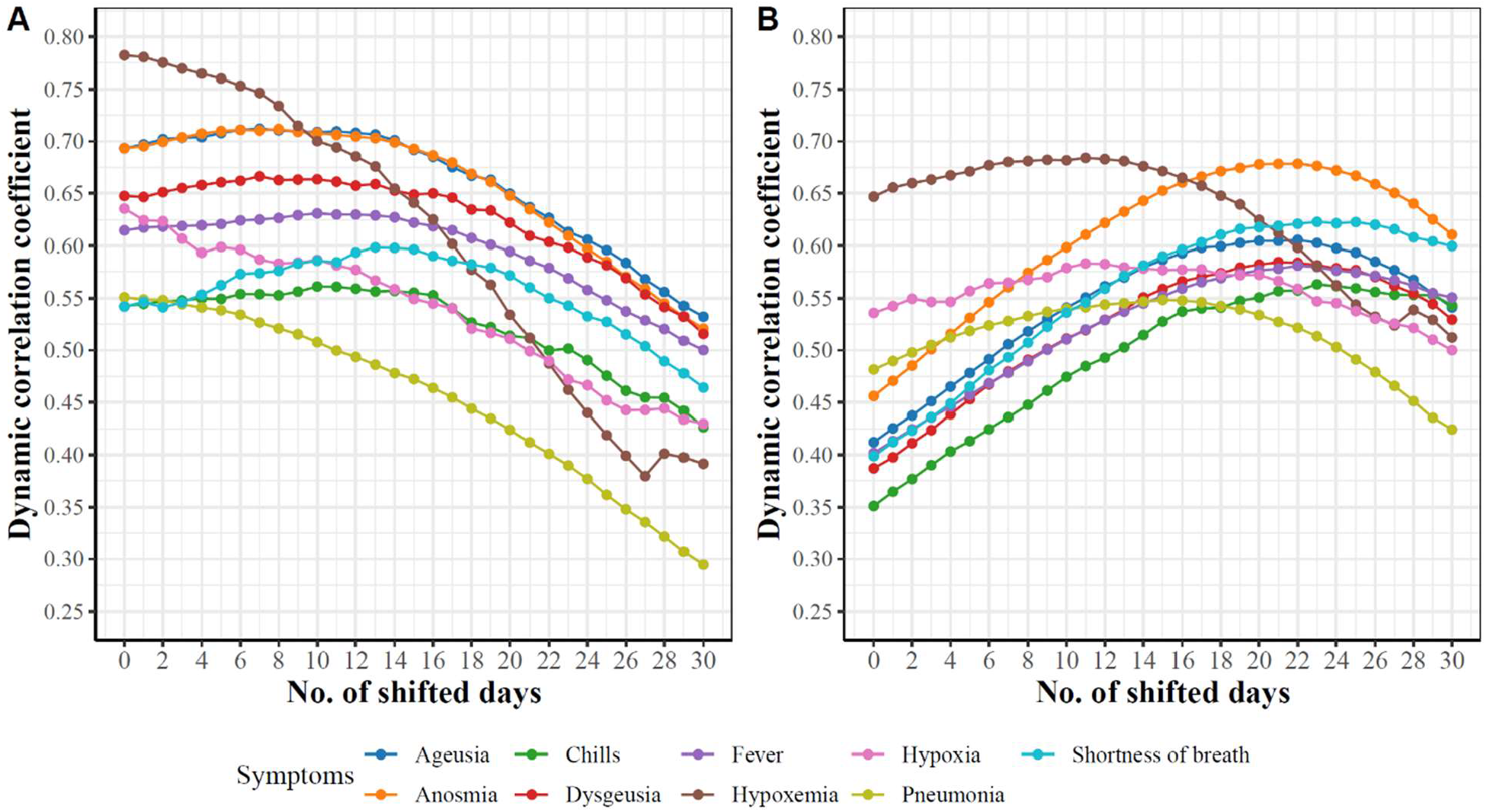
Dynamicity of the pairwise associations between the trajectories of nine symptoms and k-days ahead trajectories of COVID-19 cases (left) and deaths (right).

For correlations with COVID-19 death cases, we noted consistent increase in the correlation coefficients for all nine symptoms until each symptom reaches its maximum value at *k* ∈ [13 − 23] before the correlations start to drop. Interestingly, we noted that the correlations for hypoxemia, hypoxia, and pneumonia started to drop around *k* = 15 days, while the correlations for other symptoms started to drop around *k* = 23.

### Exploratory functional data analysis of Google search trends for hypoxemia

Since hypoxemia is the most correlated symptom with both COVID-19 confirmed and death functional data, we report an exploratory FDA of the Google daily search trends for hypoxemia. In addition, we included the results when this exploratory analysis was applied to anosmia and shortness of breath in Supplementary Figs. S3-S12.

First, Fig. 15 (left) shows the smoothed curves for the 51 states. The mean curve had three waves in the searches for hypoxemia. The first wave peaked on April 6^th^ where NY, NJ DC, and CT were the leading states with the largest observed daily normalized searches for hypoxemia. The second wave peaked in mid-July with three leading states, AZ, TX, and FL. The third wave peaked on October 5^th^ and the top three leading states were HI, MT, and AZ. Overall, these trajectories identified the three waves of COVID-19 spread and some of the leading states in each wave. Fig. 15 (right) shows the scatter plot of the 51 states in the two-dimensional space defined by the top FPC scores counting for 76% of the variability in the curves. We noted that the four leading states in the first wave had the largest second FPC scores as well as positive first FPC scores. The leading states in the second wave were located in the fourth quadrant (positive FPC1 scores and negative FPC2 scores). The figure also shows four outlier states with negative FPC2 scores, VT, WY, AK, and ND. The trajectories of these four states as well as the mean curve for the 51 states are provided in Supplementary Fig. S13. We observed that VT was a clear outlier as its curve was far below the mean curve during the time interval spanning mid-May to mid-September. The remaining three curves were the farthest curves below the mean curve from the study starting date until mid-September.

**Fig. 15.**
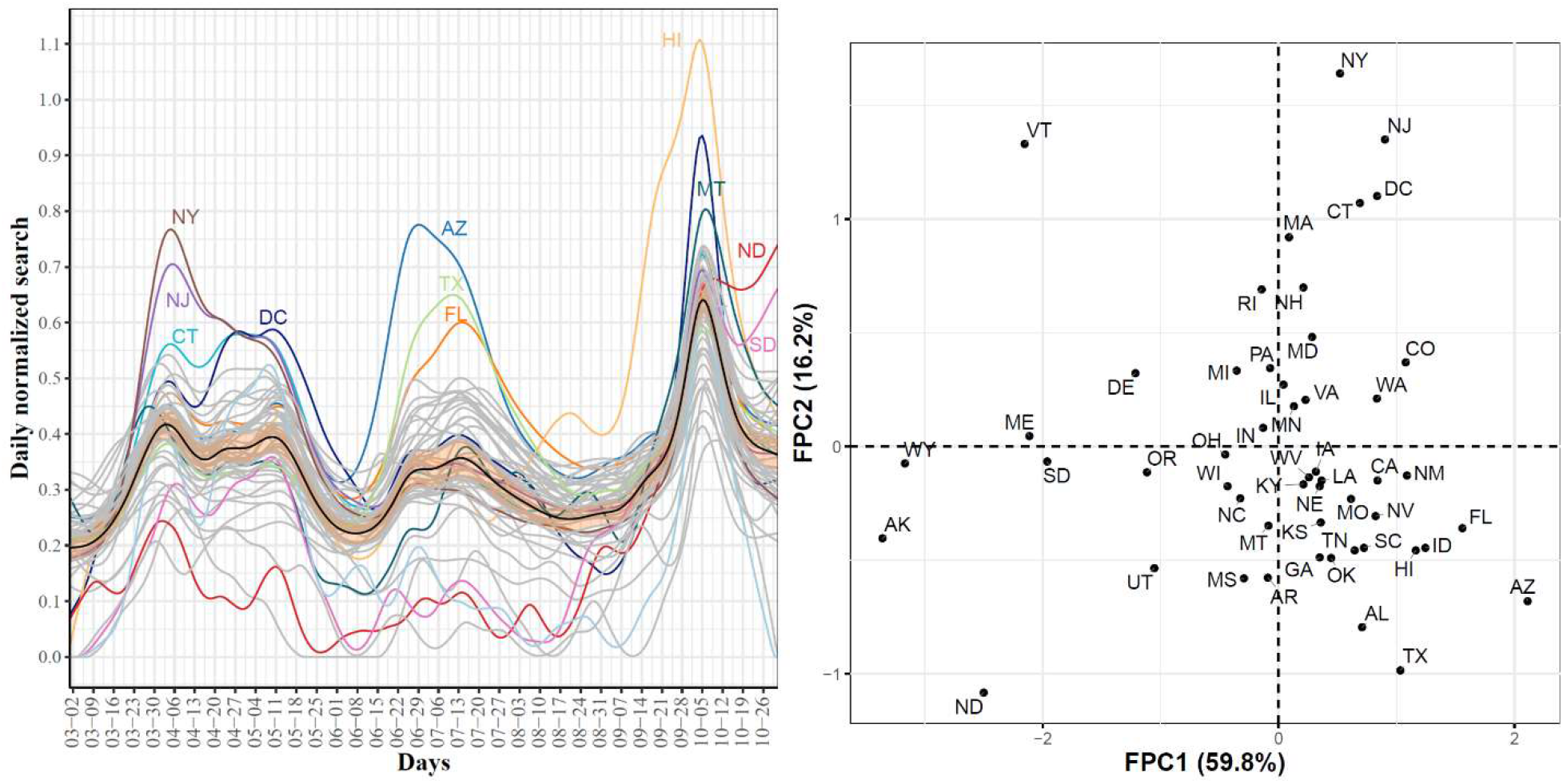
State-level trajectories of the Google daily searches for hypoxemia and their projections into a two-dimensional space determined using the first two FPC scores. The mean curve is highlighted in black and the orange ribbon corresponds to the 95% confidence band.

Second, Fig. 16 shows the first four eigenfunctions accounting for 91.1% of the total variability in the hypoxemia trajectories. Despite the overall high dynamic correlation coefficient between the trajectories of COVID-19 confirmed cases and Google searches for hypoxemia, we noted that the four eigenfunctions summarizing the modes of variations in the two datasets were different from those summarizing trajectories of the COVID-19 confirmed cases (See Figures. 3 and 16). The same observation is also valid when comparing the scattered plots of the two datasets in the two-dimensional FPC scores (See Figs. 4 and 16 (right)). To investigate the modes of variability in these two datasets, we report the results of applying the functional correlation analysis to the two datasets in the following paragraph.

**Fig. 16.**
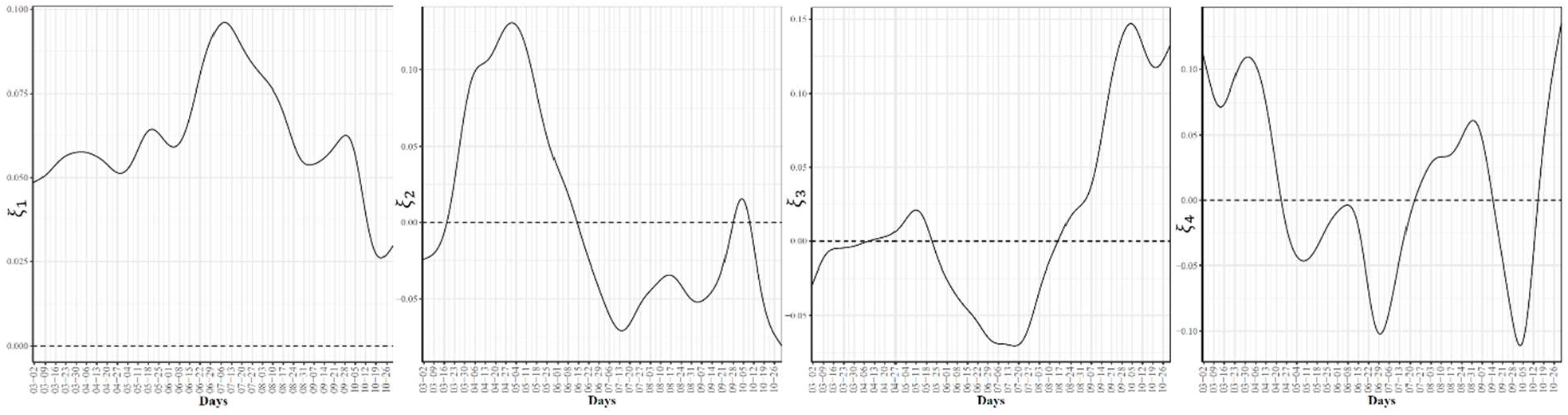
First four eigenfunctions of the state-level trajectories for the Google daily searches for hypoxemia.

Third, Fig. 17 shows the two-dimensional scatter plots of the first and second canonical scores of the COVID-19 confirmed cases and Google searches for hypoxemia sets of curves, respectively. The correlations for the first and second canonicals were 0.99 and 0.98, respectively. The scattered plot of the 51 states in the two-dimensional space defined using the first canonical scores shows that the leading states in the second wave in the hypoxemia trajectories had the highest first canonical scores, while the leading states of in the third wave (except HI since it’s a leading state in the hypoxemia trajectories but not in the trajectories of the COVID-19 confirmed cases) had the lowest first canonical scores. The scattered plot of the 51 states using their second canonical scores shows that ND and SD had the highest second canonical scores followed by the states in the second wave, while the leading states in the first wave had the lowest second canonical scores.

**Fig. 17.**
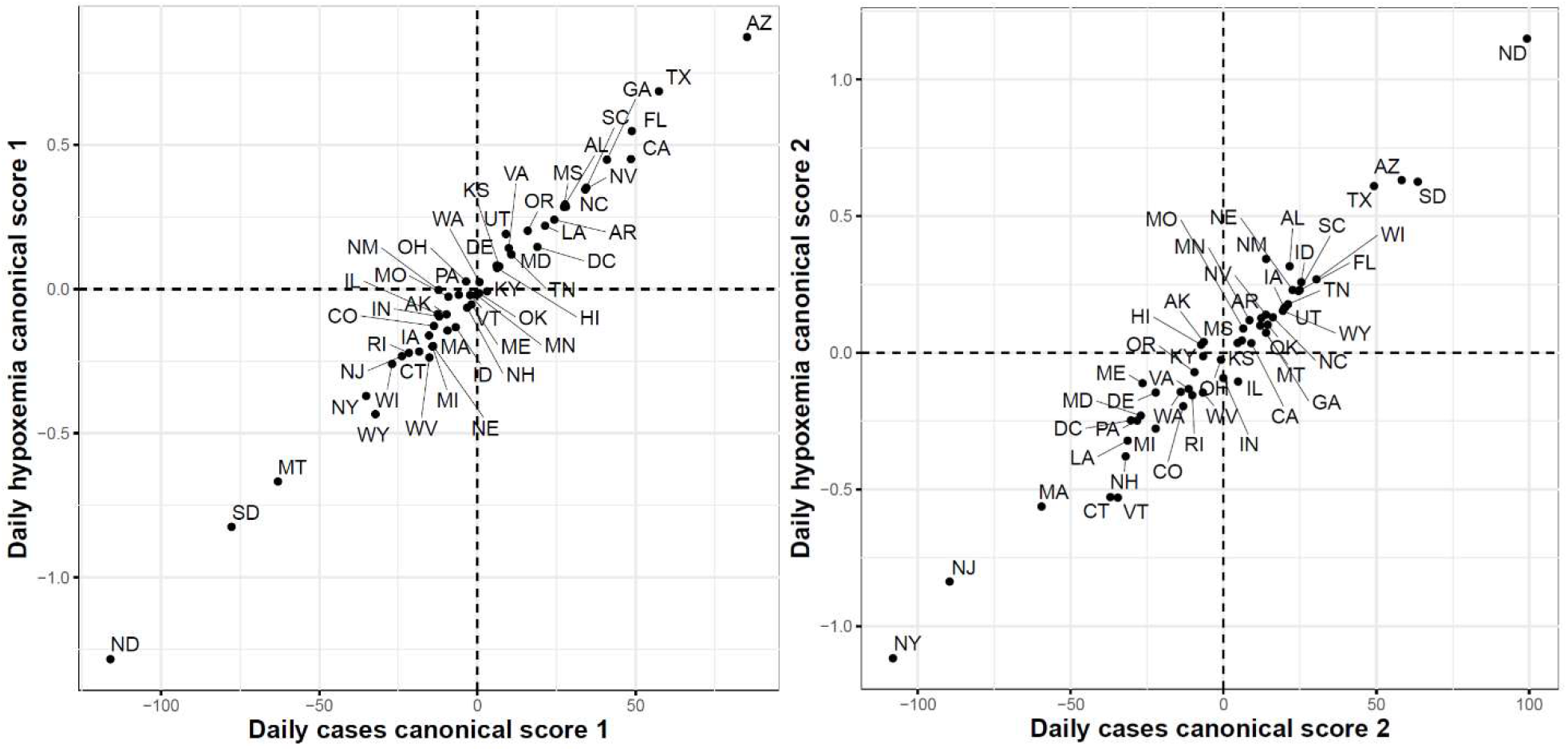
First (left) and second (right) functional canonical variables scores of the COVID-19 confirmed cases versus the Google daily searches for hypoxemia.

Finally, Fig. 18 shows the results of the FCCA when applied to COVID-19 death cases and Google searches for hypoxemia functional datasets. The correlations for the first and second canonicals were 0.96 and 0.95, respectively. The scattered plot of the 51 states using their first canonical variables shows that the states with the highest canonical scores were the states with the largest numbers of reported death cases between March and June. Interestingly the three states with the largest numbers of death cases at the end of the study period (ND, MD, and SD) had the lowest second canonical scores.

**Fig. 18.**
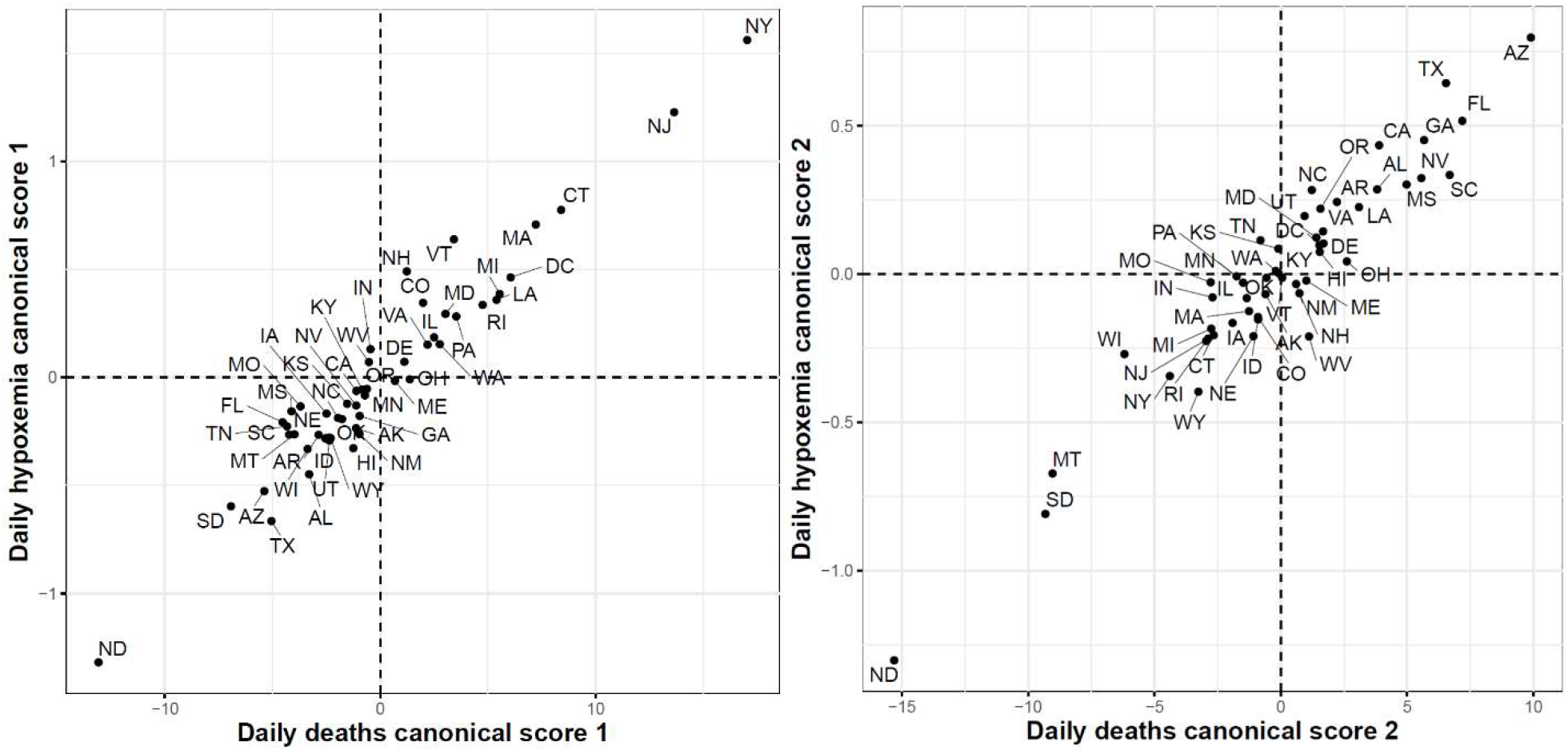
First (left) and second (right) functional canonical variables scores of the COVID-19 deaths versus. the Google daily searches for hypoxemia.

## Discussion

In this study, we conducted functional data analysis of three time-series datasets: CDC COVID-19 daily confirmed cases; CDC COVID-19 daily death cases; and Google COVID-19 symptoms search trends. Particularly, we utilized different functional data analysis techniques (including functional principal component analysis, dynamic correlations, and functional canonical correlation) to analyze and categorize different patterns of the time-dynamics of COVID-19 confirmed and death cases and to identify associations between trajectories of Google COVID-19 symptoms search trends and COVID-19 trajectories of confirmed and death cases.

Analysis of the state-level trajectories of the data demonstrated variations in the time-dynamics of COVID-19 confirmed and death cases as well as Google search trends for COVID-19 related symptoms such as hypoxemia, ageusia, and anosmia. Visualization of the trajectories revealed three waves of the spread of COVID-19 in the United States. These three waves were noted not only in the COVID-19 confirmed case trajectories but also death trajectories and the trajectories of Google searches for symptoms related to COVID-19 infection.

Although the virus spread faster during the second and third waves, we noted that the death rates were the highest during the first wave of the pandemic. The reasons are not entirely known [38] but this observation has been justified (in part) by multiple factors including our improved medical knowledge on how to better treat the COVID-19 patients, more preparedness in our health care system, some effects of the summer months, and the discovery of many mild or asymptomatic COVID-19 cases in later waves compared to the first wave when testing was often restricted to the sickest individuals. Other potential factors that haven’t been yet verified include: the virus became less deadly, vulnerable people became more protected (e.g., using social distancing and masks).

FPCA is a widely used and powerful tool in FDA. FPCA is critical for exploratory functional data analysis, dimensionality reduction, functional clustering, functional classification, and functional linear regression [39]. In this work, we found that the first two principal components can reliably identify the outlier states (i.e., the leading states in the three COVID-19 waves observed in the trajectories data). Moreover, we used functional eigenvalue analysis to explore modes of variations in the data. Finally, we used the leading four FPC scores to model each curve in a reduced dimensional space and applied the *k*-means algorithm to identify groups of states with common functional behavior. Our cluster analysis results suggested that there were seven different patterns of the COVID-19 spread across the 51 states (See Fig. 10). On the other hand, we found fewer variations across the 51 states when their death trajectories are clustered. Only five disjoint groups were identified and one of them includes a single state, NY.

All functional data analyses, discussed in the preceding paragraphs, are applicable to a single functional dataset. To explore the associations between two functional datasets (i.e., Google COVID-19 search trends for symptoms and COVID-19 confirmed/death cases), we utilized dynamic correlation and functional canonical correlation analysis. First, the dynamic correlation was used to quantify significant associations between each of the 422 symptoms functional datasets and COVID-19 confirmed/death cases functional datasets. Our results revealed two discrepancies between COVID-19 confirmed and death cases data related to their associations with Google COVID-19 search trends for symptoms: i) Google COVID-19 search trends for symptoms had stronger correlations with COVID-19 confirmed data than COVID-19 death data; ii) Dynamics of the associations between Google COVID-19 search trends for symptoms and COVID-19 confirmed cases were different from those of the associations between Google COVID-19 search trends for symptoms and COVID-19 death cases. Second, functional canonical correlation analysis (FCCA) was utilized to explore the relationships between Google search trends for hypoxemia and COVID-19 confirmed/death cases functional datasets. Our FCCA results acknowledged our results obtained using dynamic correlation analysis that suggests stronger association of Google search trends for hypoxemia and COVID-19 confirmed cases than COVID-19 death cases obtained using dynamic correlation and demonstrated the existence of common shared two-dimensional spaces between these pairs of functional datasets. Overall, our results demonstrated strong associations between Google search trends for several COVID-19 related symptoms and COVID-19 confirmed and death cases in the US.

The present study has some limitations. First, our clustering analyses have identified different patterns from the trajectories of COVID-19 confirmed and death cases in the US from the beginning of the pandemic until the end of October 2020. However, these patterns are dynamic and might change after extending the study time interval. Second, the numbers of daily positive COVID-19 cases reported during the first wave are underestimated due to insufficient testing in most of the states during the first months of the pandemic. Third, our analyses have focused on exploring the associations between Google search trends and COVID-19 spread and mortality trajectories while ignoring other potential factors such as mobility [40, 41] and environmental factors [42, 43] (e.g., temperature and humidity).

## Conclusion

We have conducted exploratory functional data analysis on daily state-level CDC COVID-19 confirmed cases and deaths time-series as well as selected Google search trends for symptoms related to COVID-19 infection. Our functional clustering results of the CDC COVID-19 spread trajectories assigned the 51 US states to seven disjoint groups each with a distinct spread pattern. Clustering of the COVID-19 death trajectories identified five distinct patterns. We have also quantified the associations between Google search trends for 422 symptoms and CDC COVID-19 trajectories for confirmed as well as death cases using dynamic correlation. Moreover, we have explored the dynamics of associations between nine top Google search trends and future COVID-19 confirmed and death cases, respectively. Finally, we have applied exploratory functional data analysis to the Google search trends for three COVID-19 related symptoms (hypoxemia, anosmia, and shortness of breath) and quantified the associations between their trajectories and the trajectories of COVID-19 confirmed and death cases using functional canonical correlation analysis. Our results and analysis framework set the stage for the development of predictive models for predicting future COVID-19 confirmed cases and deaths using historical data and Google search trends for identified symptoms, which is the focus of our ongoing work.

## Data Availability

The numbers of daily COVID-19 confirmed cases and deaths were obtained from the Centers for Disease Control and Prevention (CDC) at https://data.cdc.gov/Case-Surveillance/United-States-COVID-19-Cases-and-Deaths-by-State-o/9mfq-cb36. The Google COVID-19 Search Trends Symptoms dataset is publicly available at https://github.com/google-research/open-covid-19-data/. The 2019 US Census data are available at https://www.census.gov/data.html.

## Availability of data and materials

The numbers of daily COVID-19 confirmed cases and deaths were obtained from the Centers for Disease Control and Prevention (CDC) at https://data.cdc.gov/Case-Surveillance/United-States-COVID-19-Cases-and-Deaths-by-State-o/9mfq-cb36. The Google COVID-19 Search Trends Symptoms dataset is publicly available at: https://github.com/google-research/open-covid-19-data/. The 2019 US Census data are available at https://www.census.gov/data.html.

## Supplementary Information

Additional file 1: Supplementary Tables S1-S2

Additional file 2: Supplementary Figs. S1-S13

