## Additional file 2 for "Associations Between Google Search Trends for Symptoms and COVID-19 Confirmed and Death Cases in the United States"

### Supplementary Figures

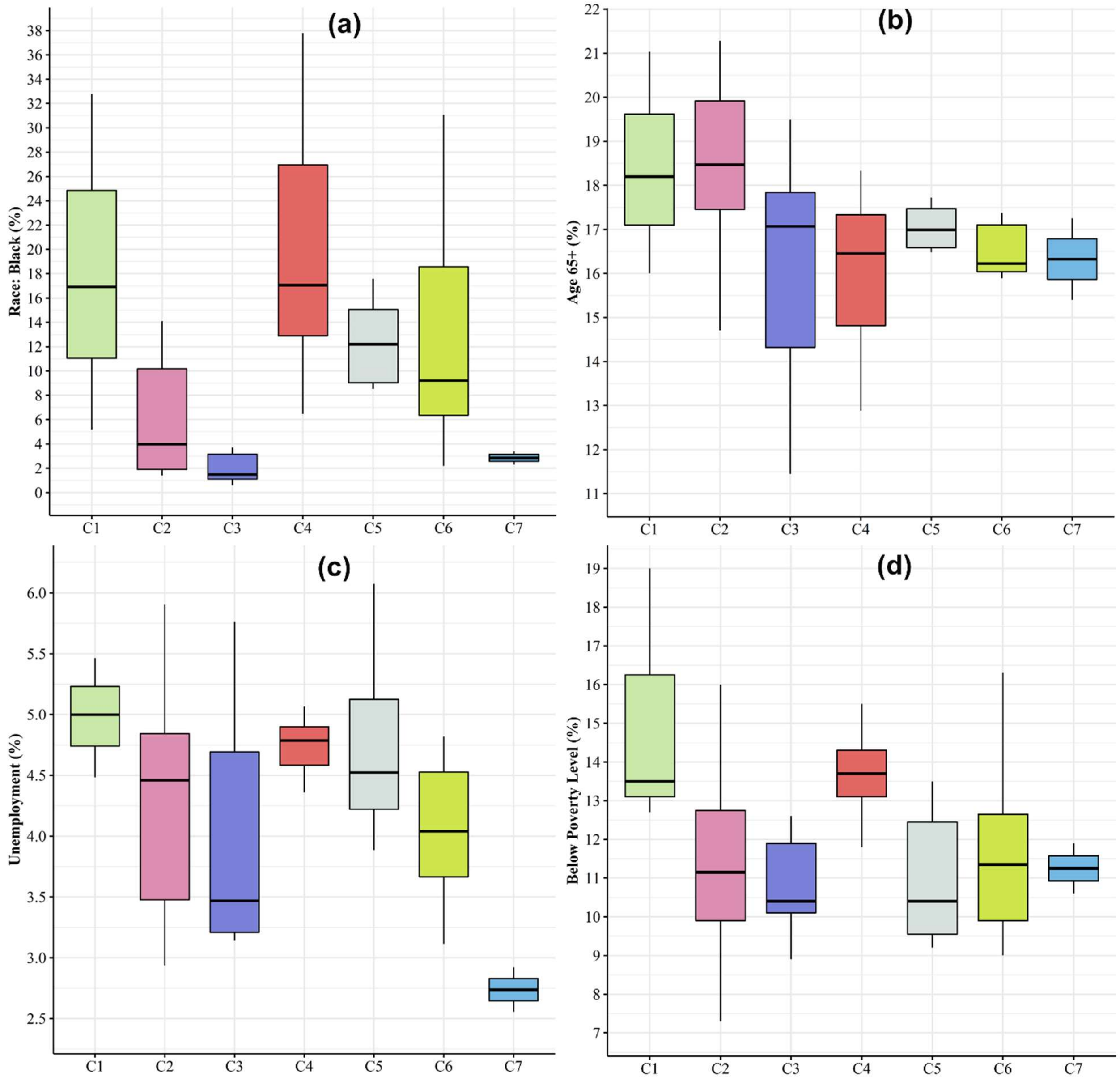


**Figure S1: Characterizing the seven groups of COVID-19 spread patterns across the US states using their boxplots of four COVID-19 risk factors corresponding to the percentage of population that are, (a) African American, (b) aged 65+ years, (c) Unemployment, (d) below the poverty level.**

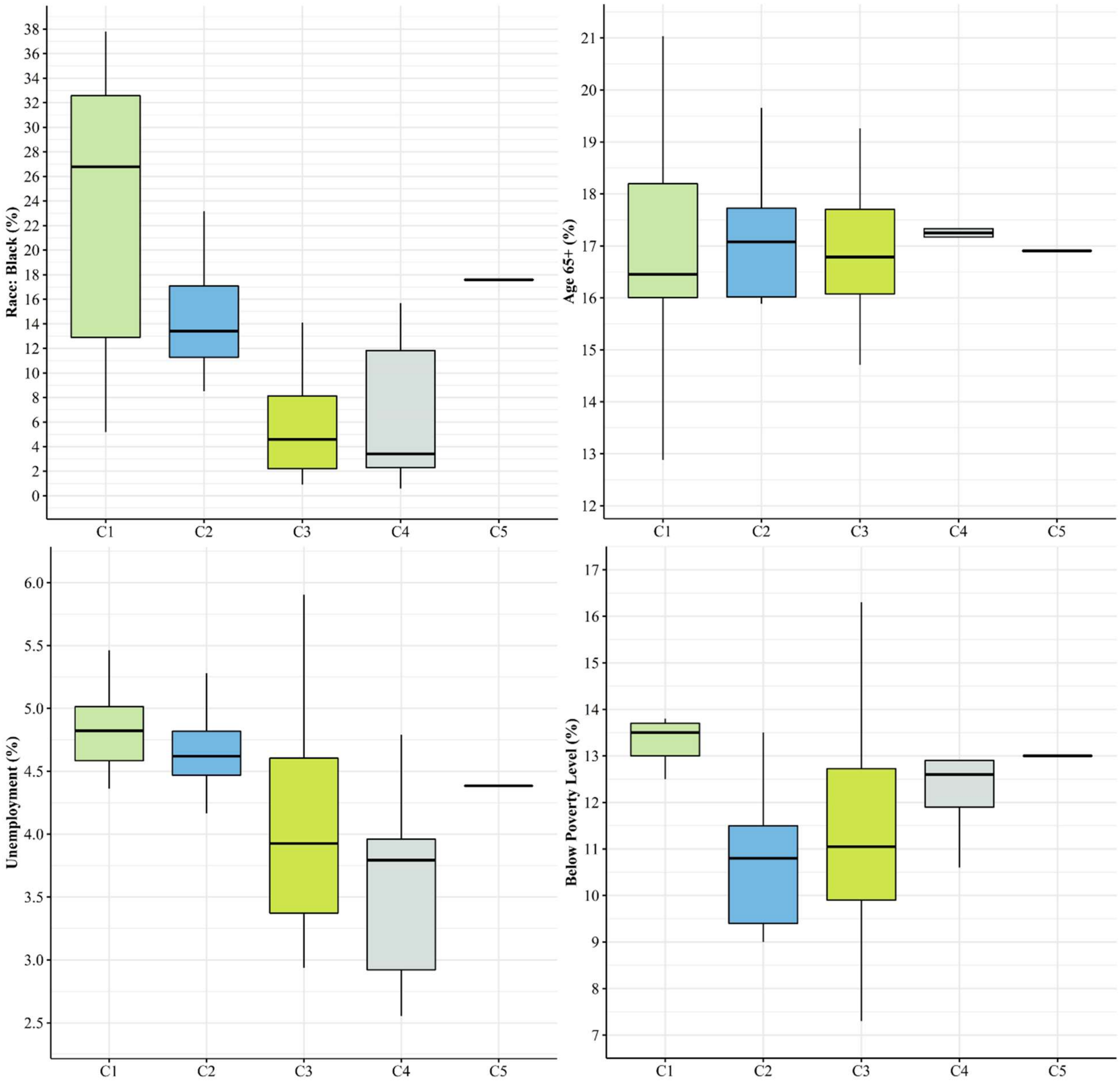


**Figure S2: Characterizing the five groups of COVID-19 death patterns across the US states using their boxplots of four COVID-19 risk factors corresponding to the percentage of population that are, (a) African American, (b) aged 65+ years, (c) Unemployment, (d) below the poverty level.**

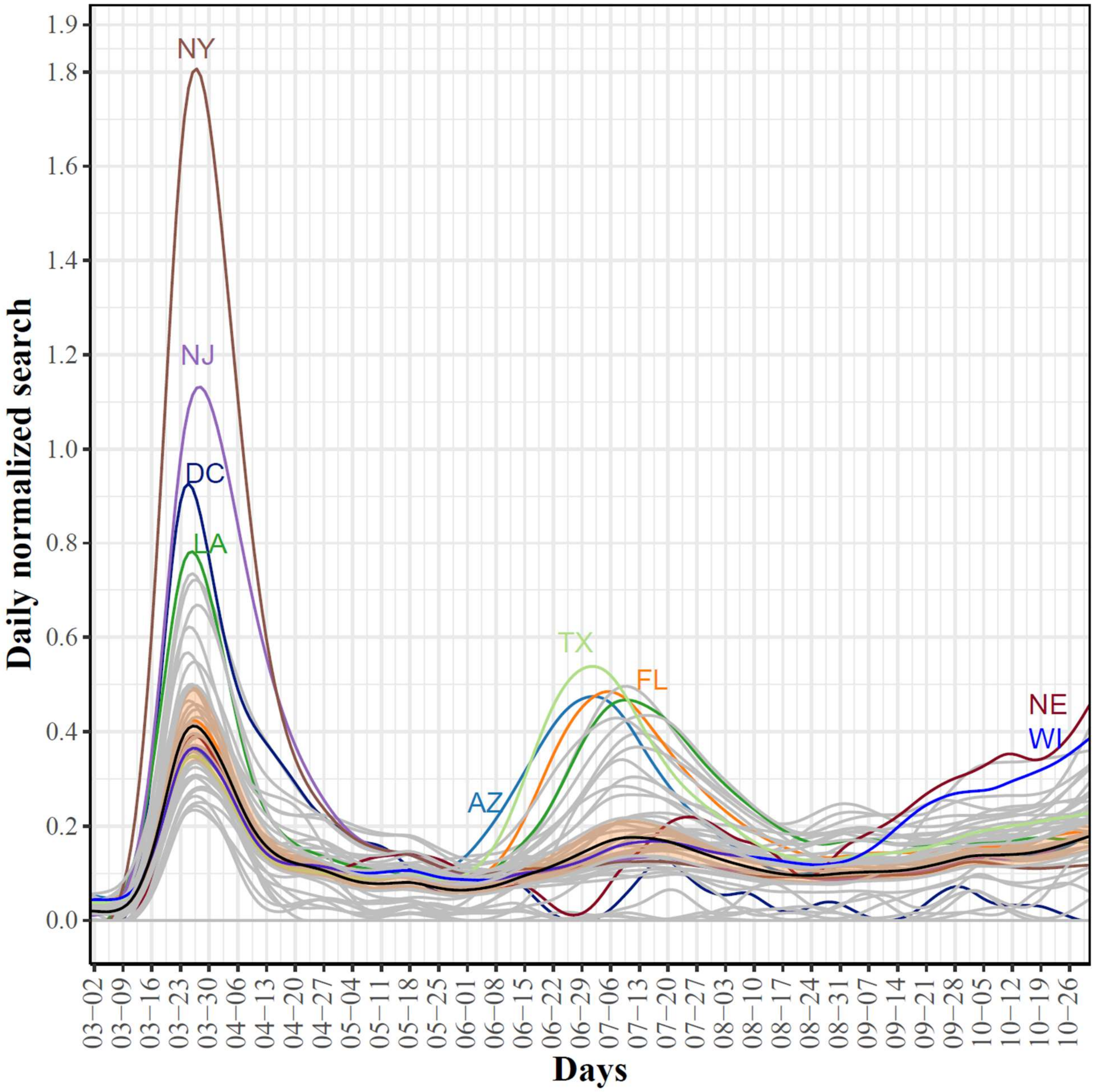


**Figure S3: State-level trajectories of the Google daily searches for anosmia. The mean curve is highlighted in black. The mean curve is highlighted in black and the orange ribbon corresponds to the 95% confidence band.**

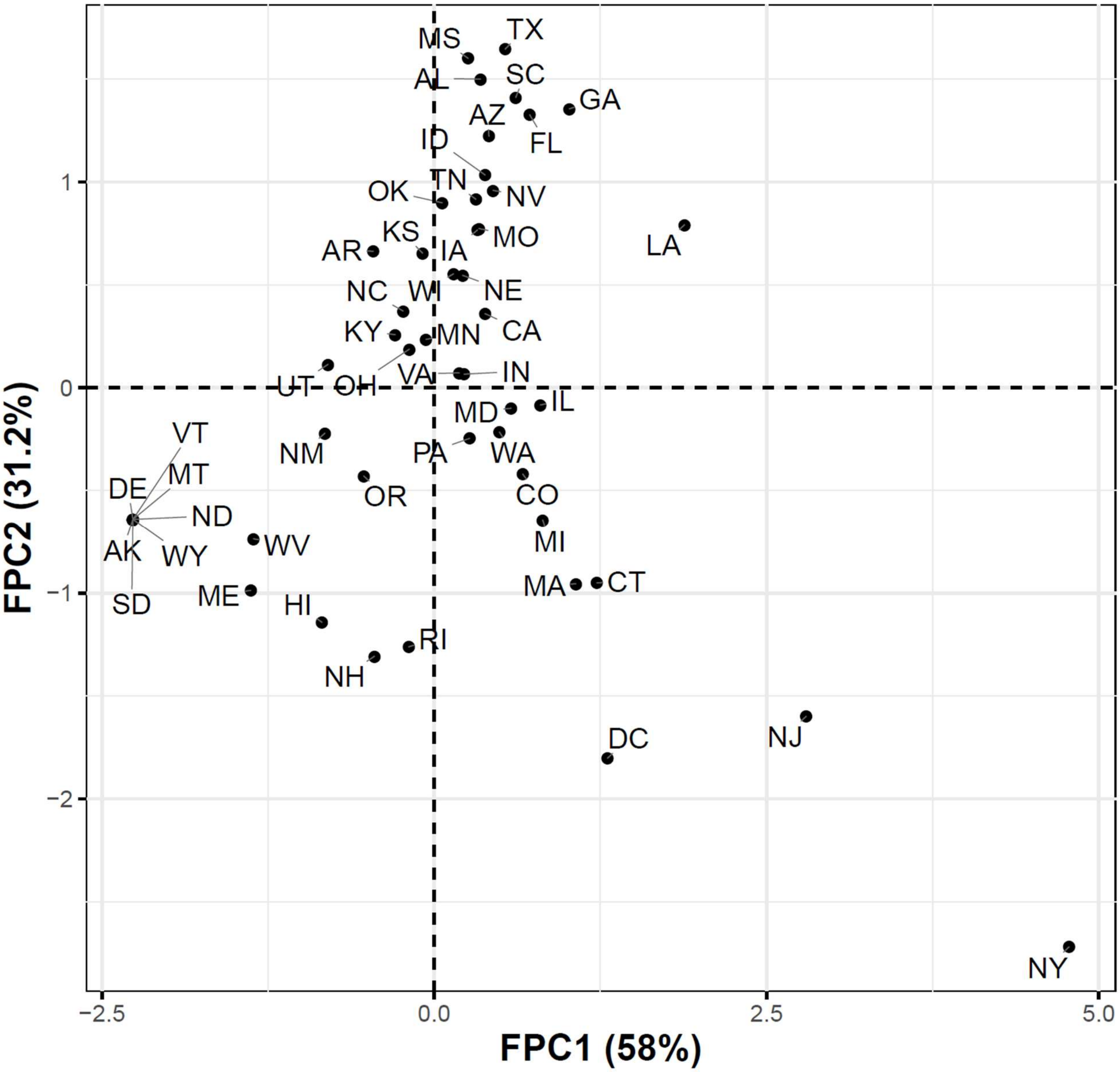


**Figure S4: Projections of the anosmia trajectories into a two-dimensional space determined using the first two FPC scores.**

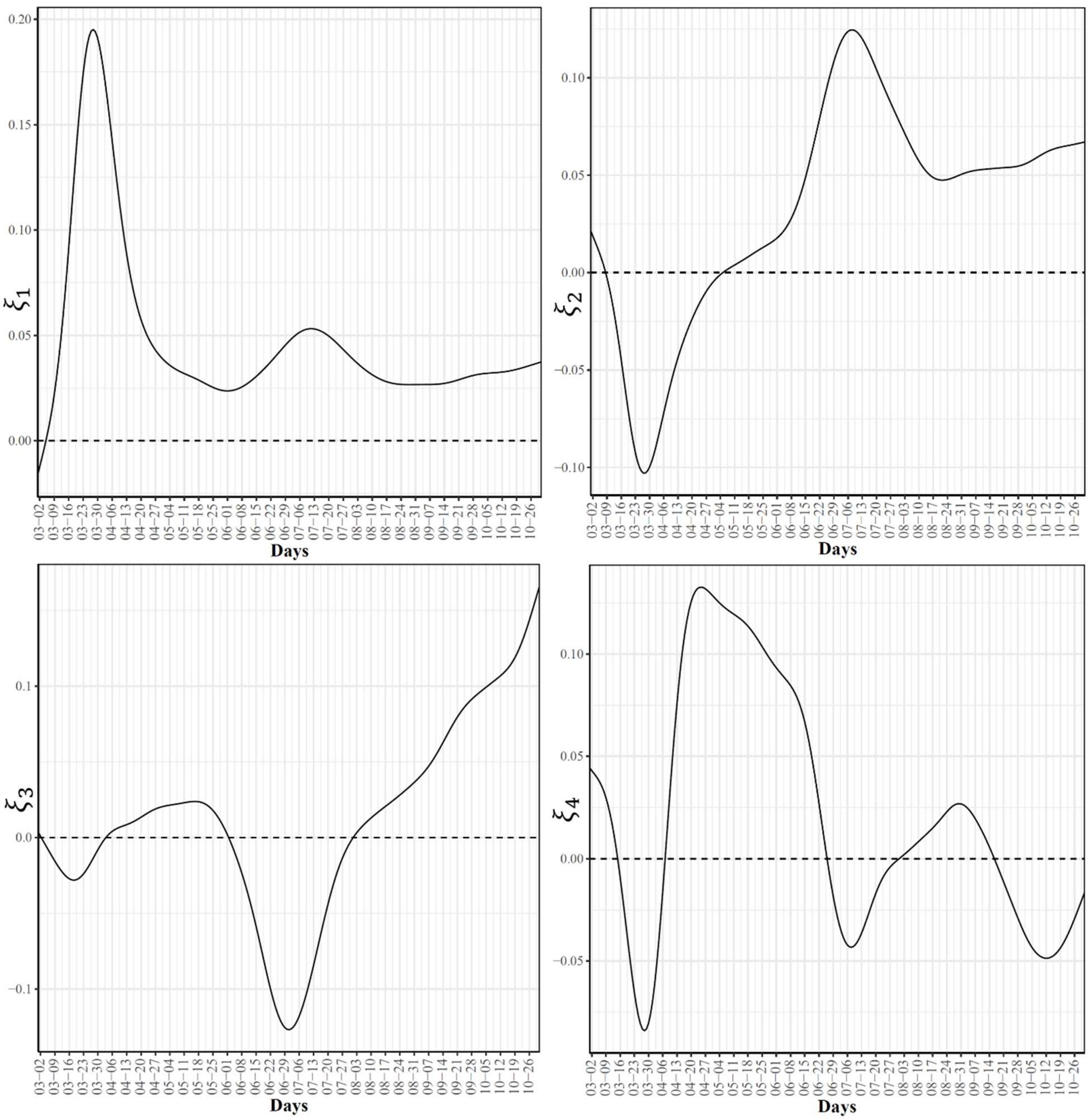


**Figure S5: First four eigenfunctions of the state-level trajectories for the Google daily searches for anosmia.**

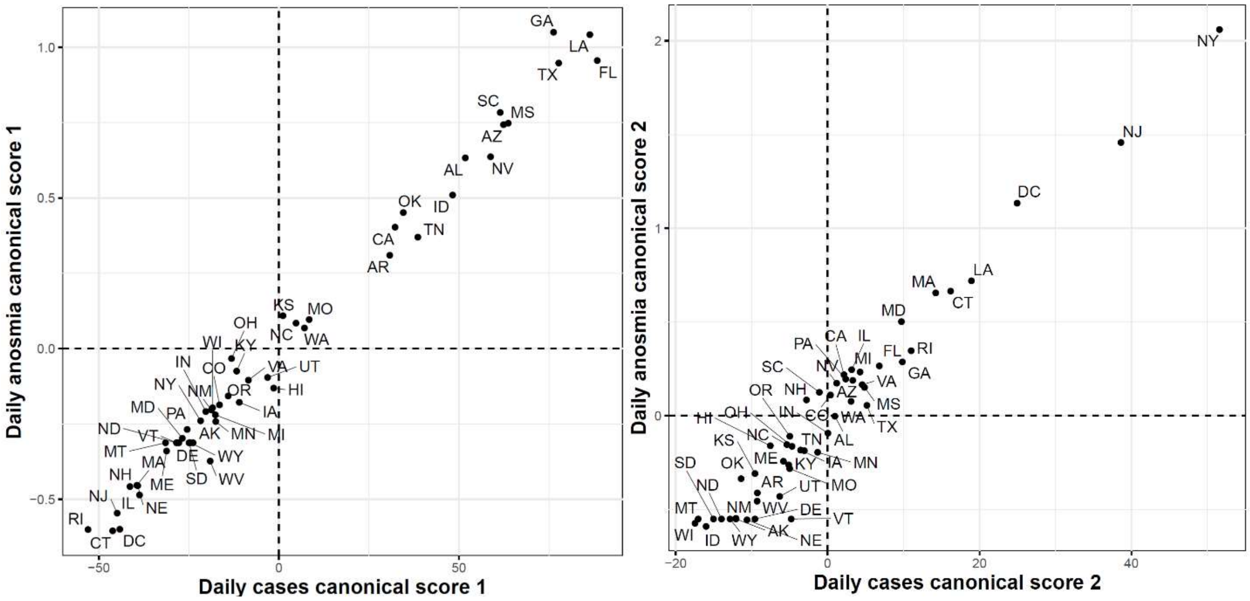


**Figure S6: First (left) and second (right) functional canonical variables scores of the COVID-19 confirmed cases versus the Google daily searches for anosmia.**

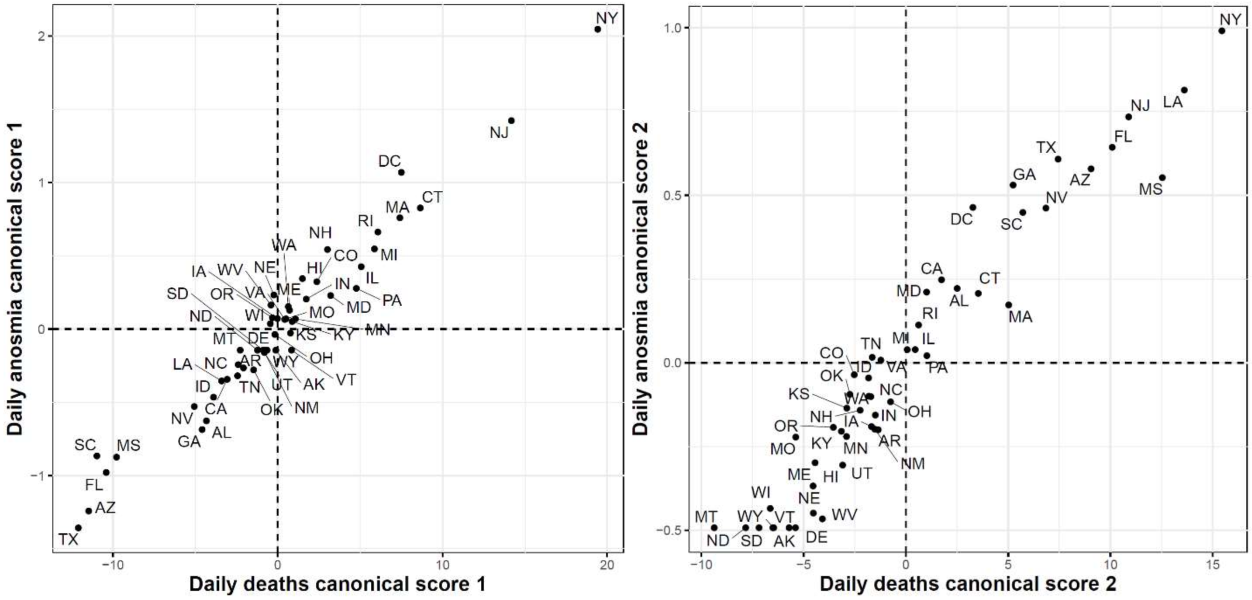


**Figure S7: First (left) and second (right) functional canonical variables scores of the COVID-19 deaths versus. the Google daily searches for anosmia.**

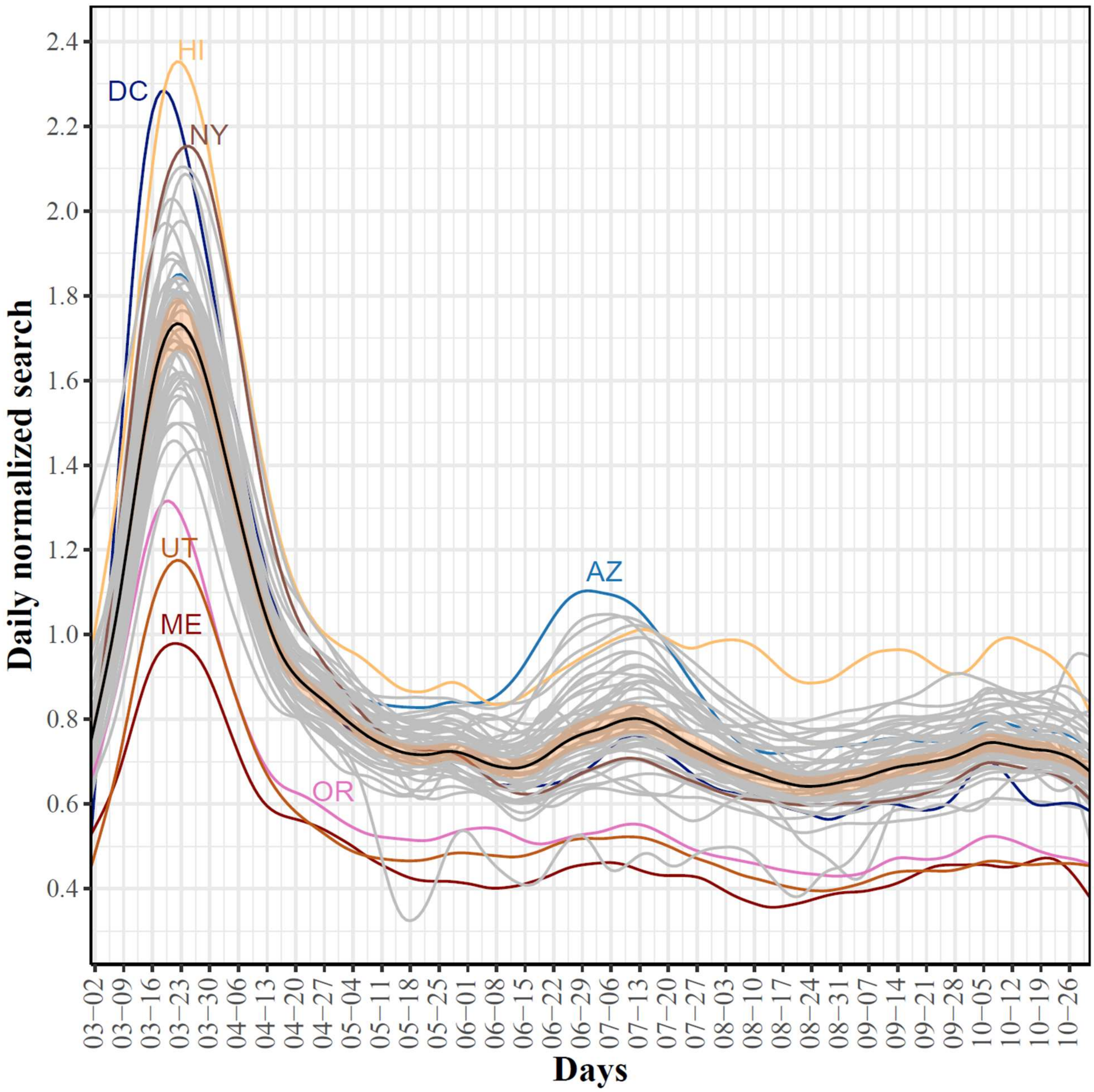


**Figure S8: State-level trajectories of the Google daily searches for "shortness of breath". The mean curve is highlighted in black and the orange ribbon corresponds to the 95% confidence band.**

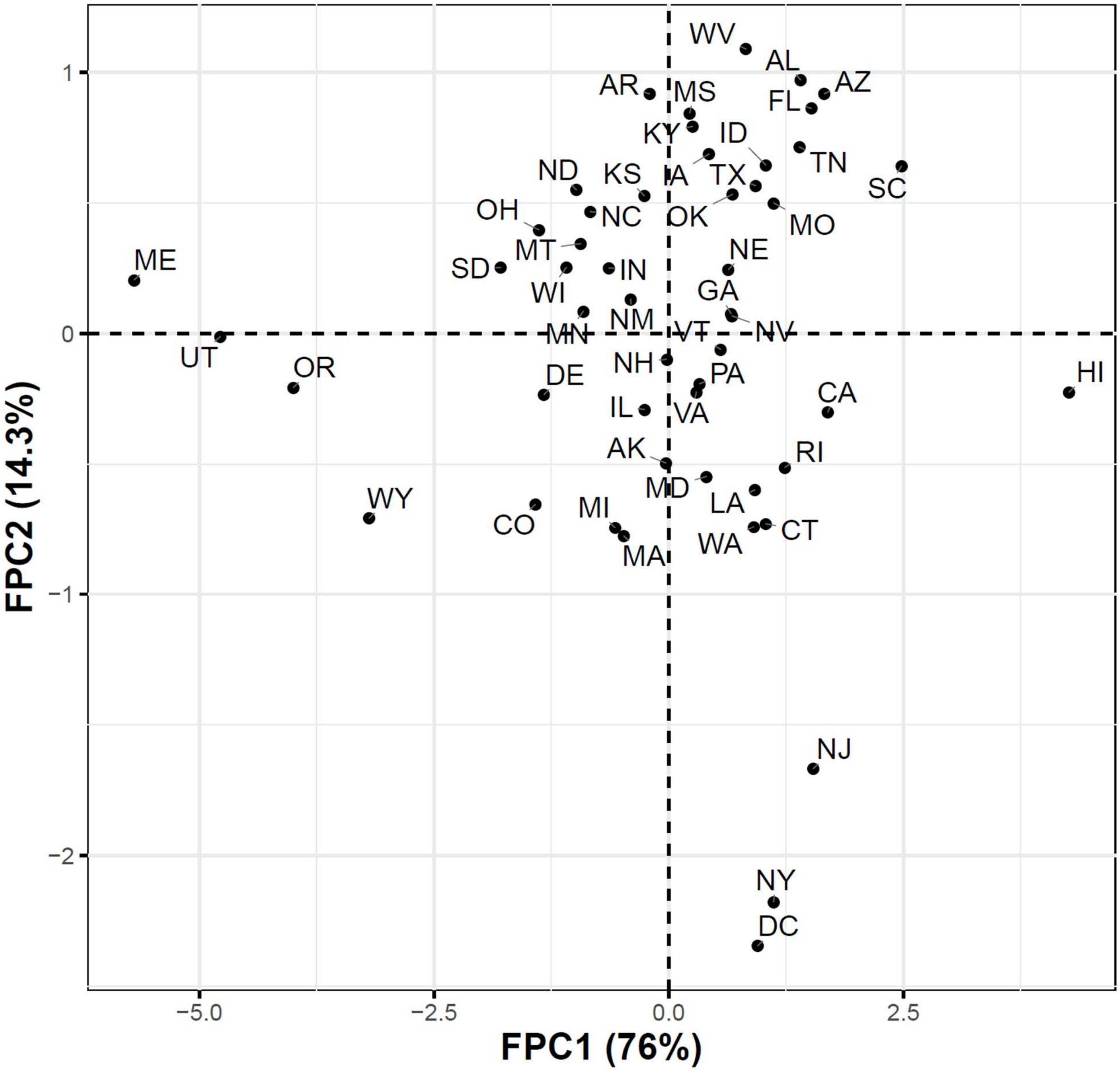


**Figure S9: Projections of the "shortness of breath" trajectories into a two-dimensional space determined using the first two FPC scores.**

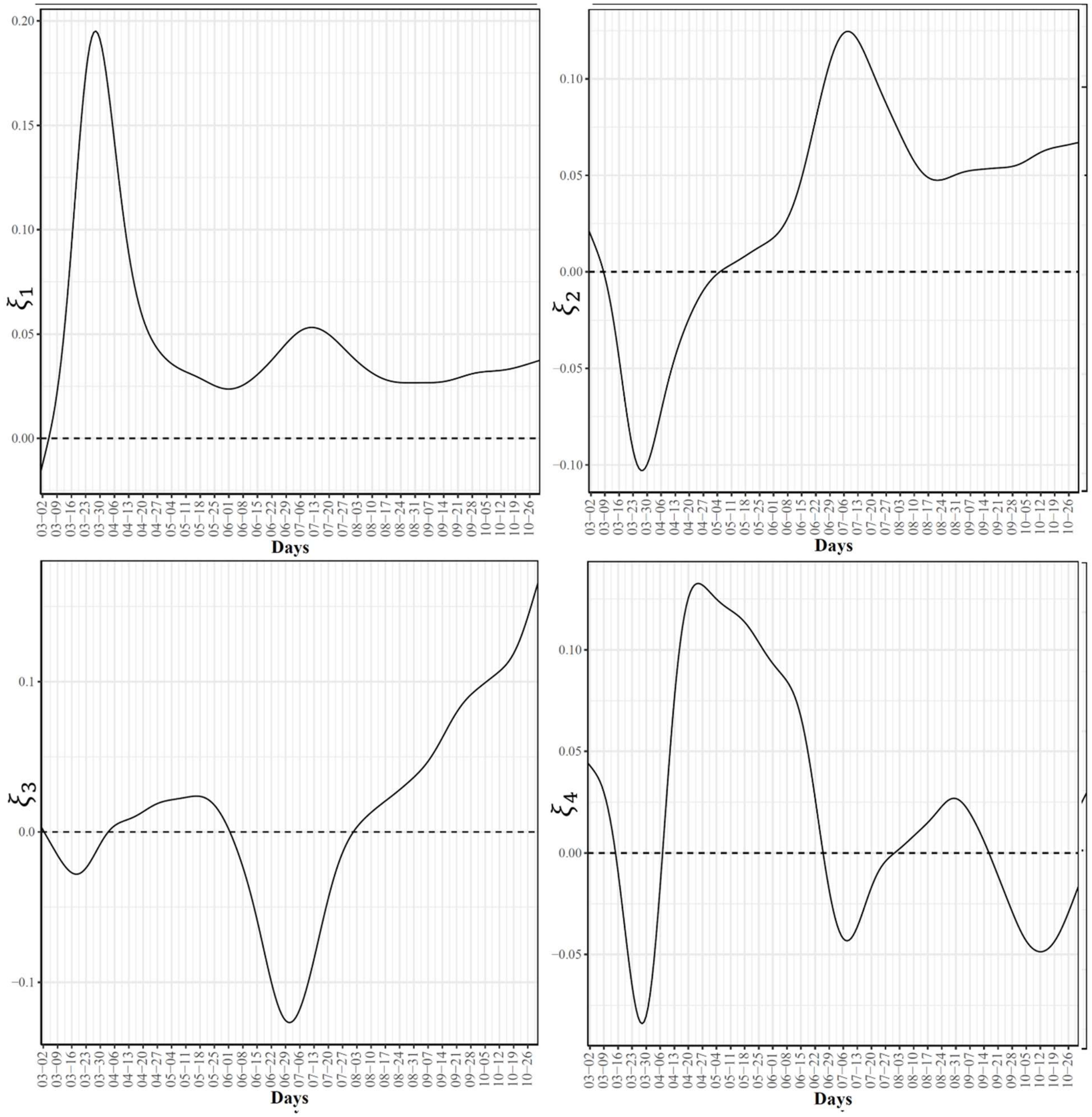


**Figure S10: First four eigenfunctions of the state-level trajectories for the Google daily searches for "shortness of breath".**

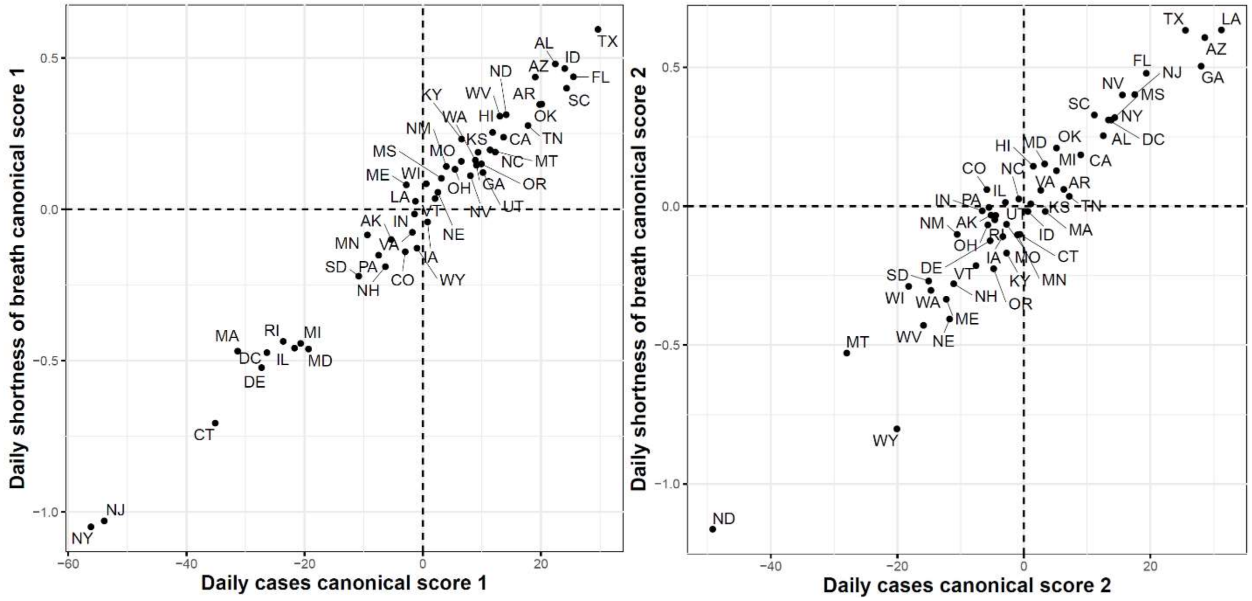


**Figure S11: First (left) and second (right) functional canonical variables scores of the COVID-19 confirmed cases versus the Google daily searches for "shortness of breath".**

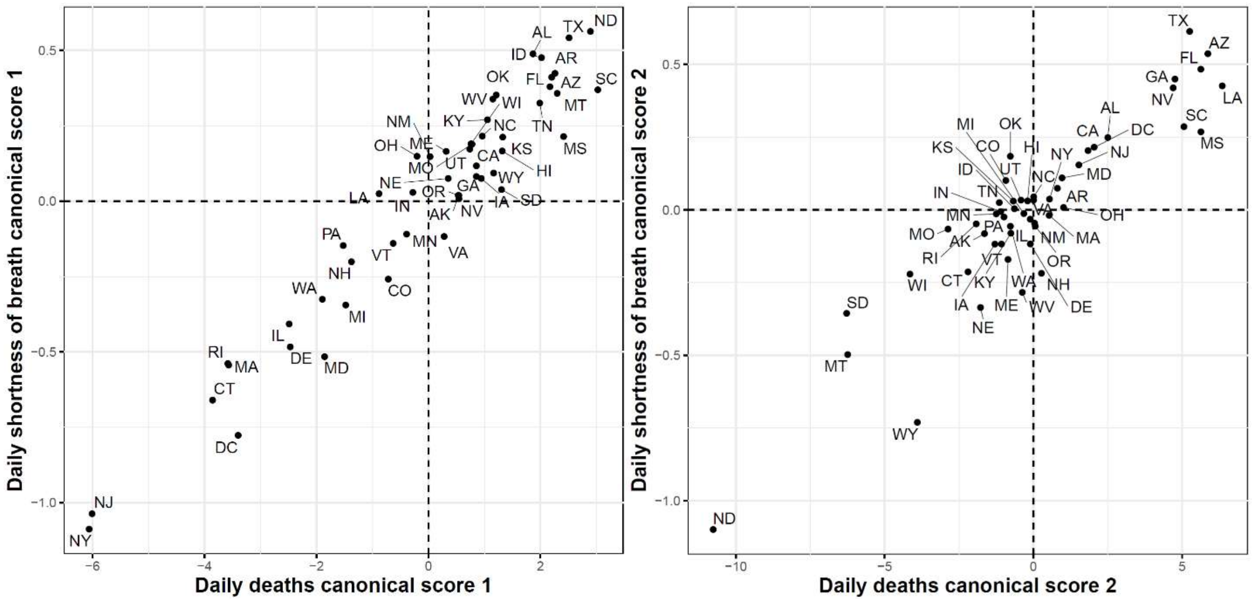


**Figure S12: First (left) and second (right) functional canonical variables scores of the COVID-19 deaths versus. the Google daily searches for "shortness of breath".**

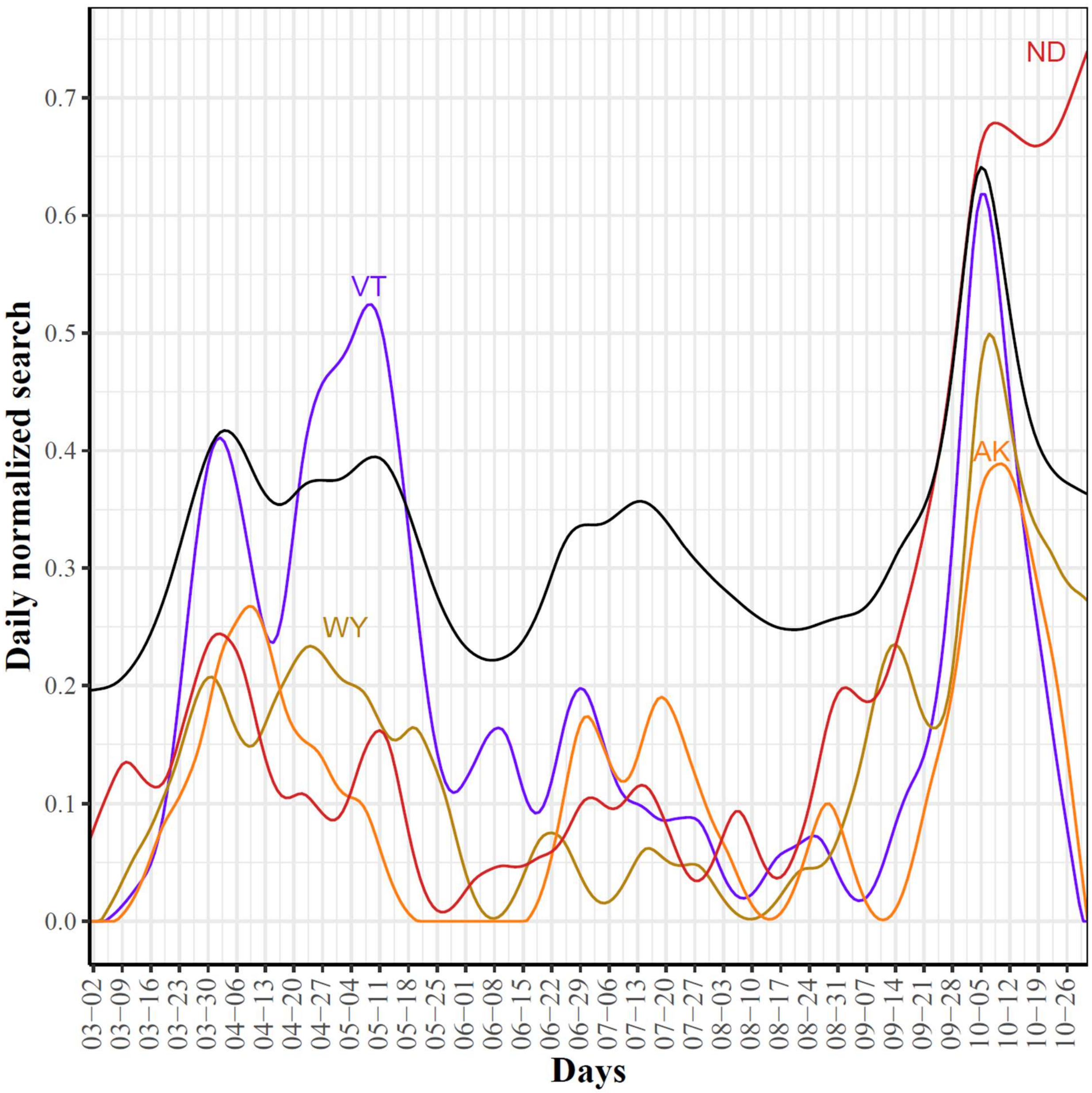


**Figure S13: Trajectories of the four outlier states with negative FPC2 scores. The black curve represents the mean curve for the 51 states.**
